# “E Pluribus Unum”: Prospective acceptability benchmarking from the Contouring Collaborative for Consensus in Radiation Oncology (C3RO) Crowdsourced Initiative for Multi-Observer Segmentation

**DOI:** 10.1101/2022.09.23.22280295

**Authors:** Diana Lin, Kareem A. Wahid, Benjamin E. Nelms, Renjie He, Mohammed A. Naser, Simon Duke, Michael V. Sherer, John P. Christodouleas, Abdallah S. R. Mohamed, Michael Cislo, James D. Murphy, Clifton D. Fuller, Erin F. Gillespie

**Author notes:** co-corresponding authors **Corresponding authors:** Clifton D. Fuller, MD, PhD, Department of Radiation Oncology, The University of Texas MD Anderson Cancer Center, 1515 Holcombe Blvd. Houston, TX, 77030, Erin F. Gillespie, MD, Department of Radiation Oncology, Memorial Sloan Kettering Cancer Center, 1275 York Ave, Box 22, New York, NY 10065. co-first authors (Diana Lin, Kareem A. Wahid).

## Abstract

**OBJECTIVE:** Contouring Collaborative for Consensus in Radiation Oncology (C3RO) is a crowdsourced challenge engaging radiation oncologists across various expertise levels in segmentation. A challenge in artificial intelligence (AI) development is the paucity of multi-expert datasets; consequently, we sought to characterize whether aggregate segmentations generated from multiple non-experts could meet or exceed recognized expert agreement.

**MATERIALS AND METHODS:** Participants who contoured ≥1 region of interest (ROI) for the breast, sarcoma, head and neck (H&N), gynecologic (GYN), or gastrointestinal (GI) challenge were identified as a non-expert or recognized expert. Cohort-specific ROIs were combined into single simultaneous truth and performance level estimation (STAPLE) consensus segmentations. STAPLE_non-expert_ ROIs were evaluated against STAPLE_expert_ contours using Dice Similarity Coefficient (DSC). The expert interobserver DSC (IODSC_expert_) was calculated as an acceptability threshold between STAPLE_non-expert_ and STAPLE_expert_. To determine the number of non-experts required to match the IODSC_expert_ for each ROI, a single consensus contour was generated using variable numbers of non-experts and then compared to the IODSC_expert_.

**RESULTS:** For all cases, the DSC for STAPLE_non-expert_ versus STAPLE_expert_ were higher than comparator expert IODSC_expert_ for most ROIs. The minimum number of non-expert segmentations needed for a consensus ROI to achieve IODSC_expert_ acceptability criteria ranged between 2-4 for breast, 3-5 for sarcoma, 3-5 for H&N, 3-5 for GYN ROIs, and 3 for GI ROIs.

**DISCUSSION AND CONCLUSION:** Multiple non-expert-generated consensus ROIs met or exceeded expert-derived acceptability thresholds. 5 non-experts could potentially generate consensus segmentations for most ROIs with performance approximating experts, suggesting non-expert segmentations as feasible cost-effective AI inputs.

## Background and Significance

Contouring, also referred to as delineation or segmentation, of regions of interest (ROIs) on medical imaging is a crucial aspect of radiation treatment planning, and has been reported as the largest single source of system uncertainty in radiotherapy [1], especially in the sense that, intrinsically, there is often no “ground truth” for absolute determination of patient-specific segmentation accuracy [2]. Manual definition and annotation of target volumes and organs-at-risk (OARs) is subject to considerable interobserver variability, even among experts [3,4], leading to inconsistent contour quality [5,6], which have been correlated with disease control decrement and increased toxicity [7–9]. Efforts to reduce manual segmentation variation have included consensus guidelines [10,11], which generally include a benchmark “gold standard” contour curated by one expert for clinical use or through a single simultaneous truth and performance level estimation (STAPLE)-consensus derived by an interdisciplinary expert panel [2]. Several studies have demonstrated that the use of contouring atlases can reduce variation in contouring[12–16], but use is limited in routine practice. More recently, Zhang et al. have shown the addition of a radiation anatomist could also reduce contour variation [17], but these efforts are still exploratory.

Auto-segmentation, broadly defined as the automated generation of contours on a digital image by a computer algorithm, has emerged as an avenue to decrease contour variability and thereby improve standardization. While automated contouring methods evolve, a significant challenge in the development of auto-segmentation algorithms is the relative paucity of curated multi-expert observer datasets sufficiently large to train machine learning models, e.g., deep learning approaches [18]. This is particularly true for disease sites such as the head and neck (H&N), which have demonstrated high interobserver segmentation variability [19,20].

As such, our team developed the Contouring Collaborative for Consensus in Radiation Oncology (C3RO), the first public crowdsourced challenge to engage radiation oncologists across various expertise levels in cloud-based image-segmentation in multiple disease sites. We sought to: 1) characterize the variability in radiation oncology segmentation performance across multiple levels of expertise compared to aggregated expert performance as “gold standard”, 2) determine whether aggregate or “composite” segmentations generated from non-experts could meet or exceed *individual* expert performance acceptability, and 3) examine the performance dynamics of consensus segmentation generation using a variable number of non-experts required to generate acceptable segmentation priors. An overview of our study is shown in **Figure 1**.

**Figure 1.**
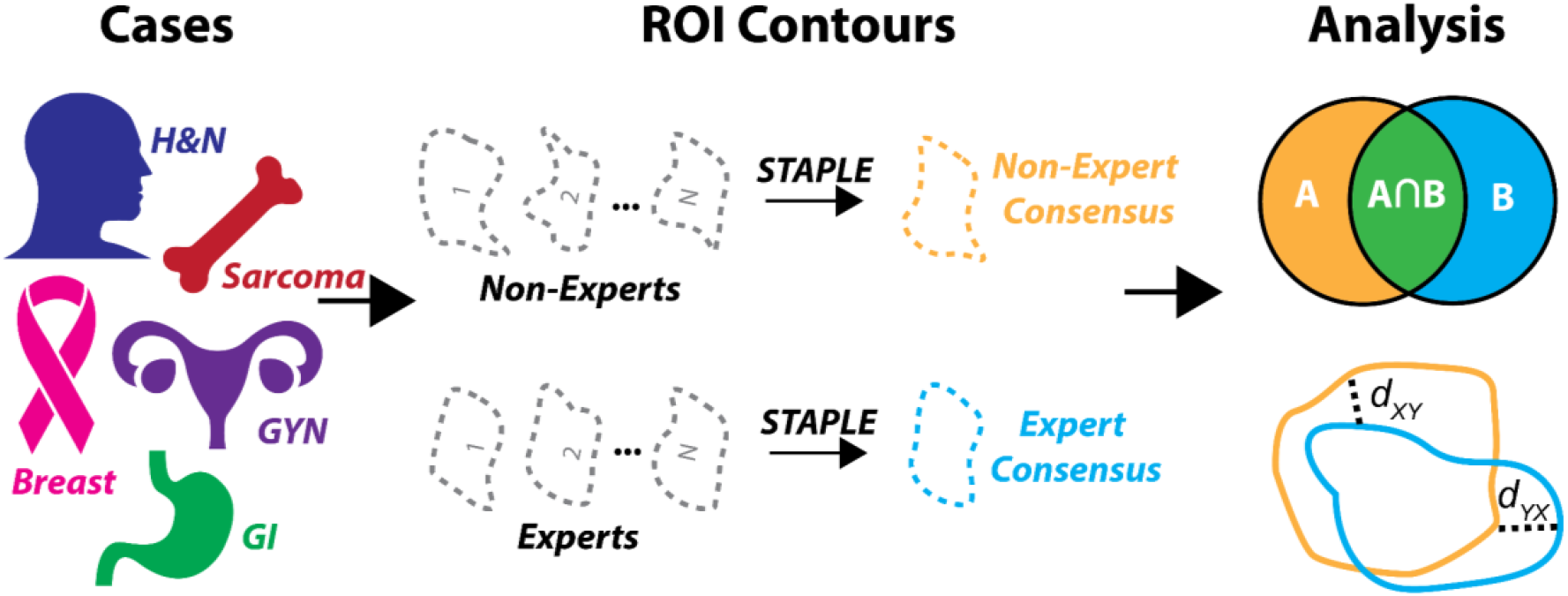
Study workflow overview. Radiotherapy planning cases across a variety of disease sites were used to crowdsource non-expert and expert region of interest (ROI) contours. These contours were then investigated to determine interobserver variability and used in consensus segmentation experiments. Abbreviations: H&N = head and neck, GYN = gynecology, GI = gastrointestinal, STAPLE = simultaneous truth and performance level estimation algorithm.

## Methods

### Study Design

C3RO, initially launched in August 2021, is an online crowdsourced challenge inviting radiation oncologists around the world to contour a new case every two months. At the end of each case, participants who have completed at least one contour are eligible to win a gift card and have access to: (1) the contours of recognized disease site experts for the case and (2) a live video podcast hosted by two to three select experts, reviewing common contouring errors and their decision-making rationale. A week after the live podcast, the recording of the expert discussion is posted on YouTube for public access (current address: https://www.youtube.com/channel/UC43Mxi5uRARSXEmSdfm6wqw, permanent archived address: https://web.archive.org/web/20220906231217/https://www.youtube.com/channel/UC43Mxi5uRARSXEmSdfm6wqw).

### Participant Recruitment

Participants were recruited through Twitter, word of mouth, the annual symposium at the Radiation Oncology Education Collaborative Study Group (ROECSG), and via eContour’s userbase. eContour is an interactive web-based platform our team developed to collect and disseminate consensus guidelines; it is now used by over 33,000 radiation oncologists from 128 countries, 12,650 of whom have been identified as practicing radiation oncologists (attending or resident) [21]. Participants were categorized as recognized experts or non-experts. Recognized experts were identified by our C3RO team (EFG, CDF, DL) based on participation in the development of national guidelines or other extensive scholarly activities and recognized expertise within the specific disease site.

### Data Collection

To register for the challenge, participants completed a baseline questionnaire that included their name, email address, affiliated institution, country, specialization, years in practice, number of disease sites treated, volume of patients treated per month for the designated tumor site, how they learned about this challenge, and reasons for participation (see **Appendix A**). Once the participant registered, they were granted access to the C3RO workspace on ProKnow (Elekta AB, Stockholm, Sweden), a cloud-based contouring platform that stores and manages the data. Completion of the baseline questionnaire served as informed consent, and the study was approved as exempt by the institutional review board at Memorial Sloan Kettering (IRB#: X19-040 A(1); approval date: May 26, 2021).

### Imaging Data

Five cases are utilized from the C3RO challenge: breast, sarcoma, H&N, gynecology (GYN), and gastrointestinal (GI). Each case contains 1 computed tomography image of a representative patient in Digital Imaging and Communications in Medicine (DICOM) format. Anonymized data for all cases were received from study collaborators. Imaging details of the cases are shown in **Appendix B**. Participants (experts and non-experts) were instructed to contour a set of representative ROIs for each case. A table of ROIs used for each case is shown in **Table 1**; ROI naming conformed to the American Association of Physicists in Medicine Task Group 263 standard [22]. Each participant generated one radiotherapy structure (RT-STRUCT) file for each ROI structure set.

**Table 1.**
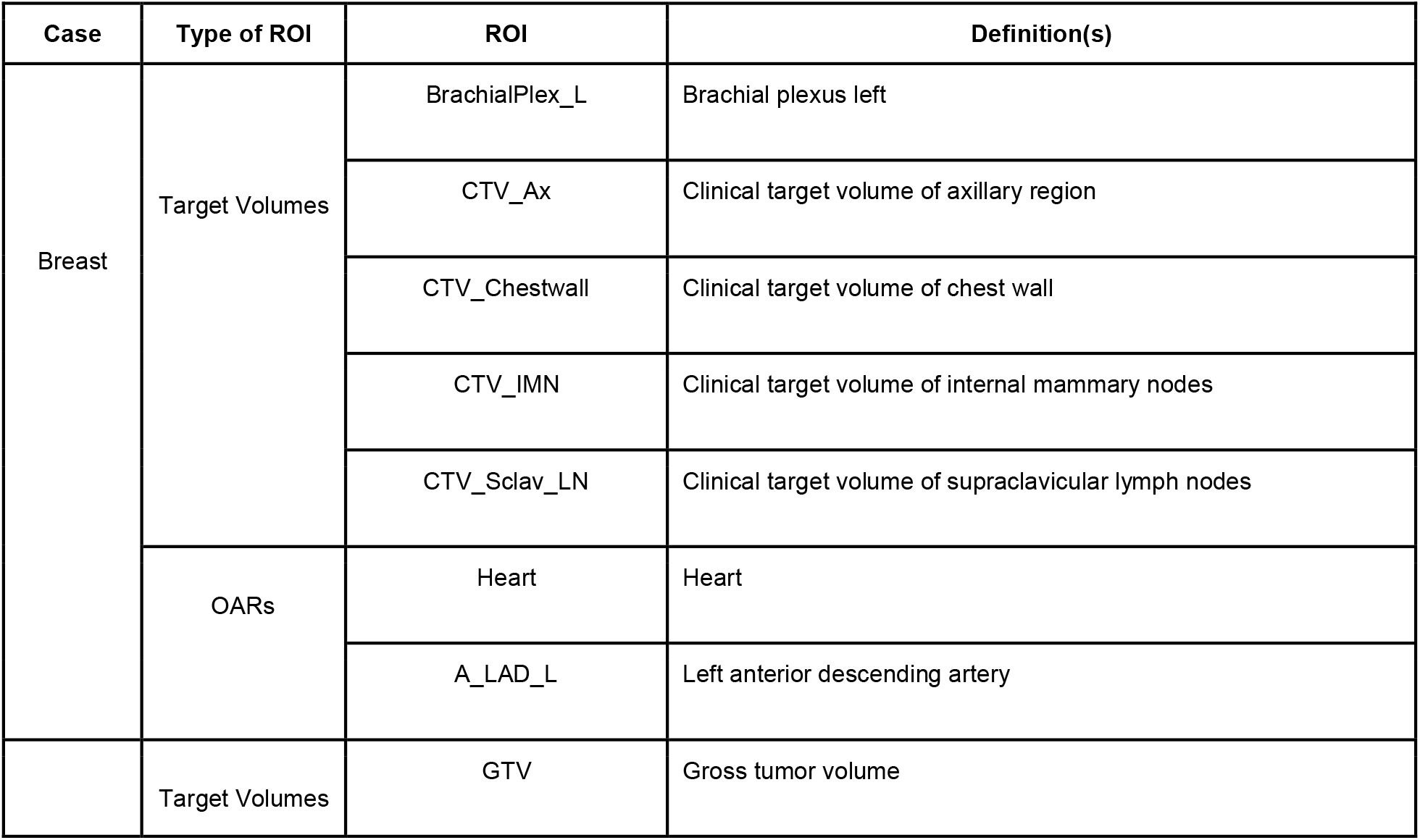
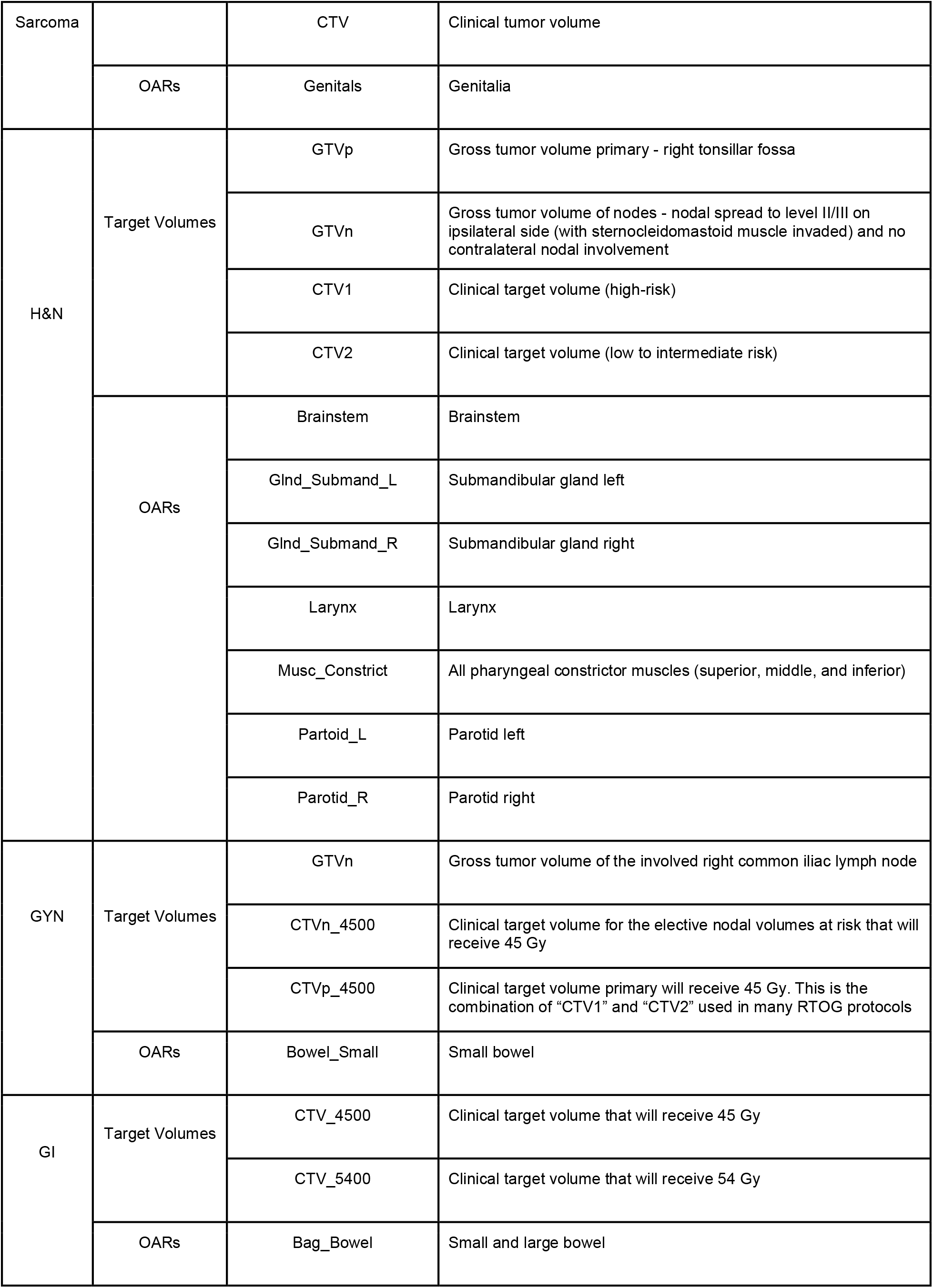
Regions of interest (ROI)s and definitions used for each case. Abbreviations: H&N = head and neck, GYN = gynecology, OARs = organs-at-risk.

### Data Processing

Images and segmentation masks were analyzed in Python v. 3.9.0. All DICOM images and DICOM RT-STRUCT files were converted to Neuroimaging Informatics Technology Initiative format using the DICOMRTTool v. 3.2.0 Python package [23].

### Consensus Methods

The simultaneous truth and performance level estimation (STAPLE) algorithm [2], a well-validated and widely implemented consensus segmentation method based on weighted probabilistic estimation, was utilized to generate consensus multi-observer ROIs for this analysis. We utilized the SimpleITK [24] STAPLE implementation with a threshold value of 0.95. An example of a consensus segmentation generated from a set of expert segmentations in **Figure 2**.

**Figure 2.**
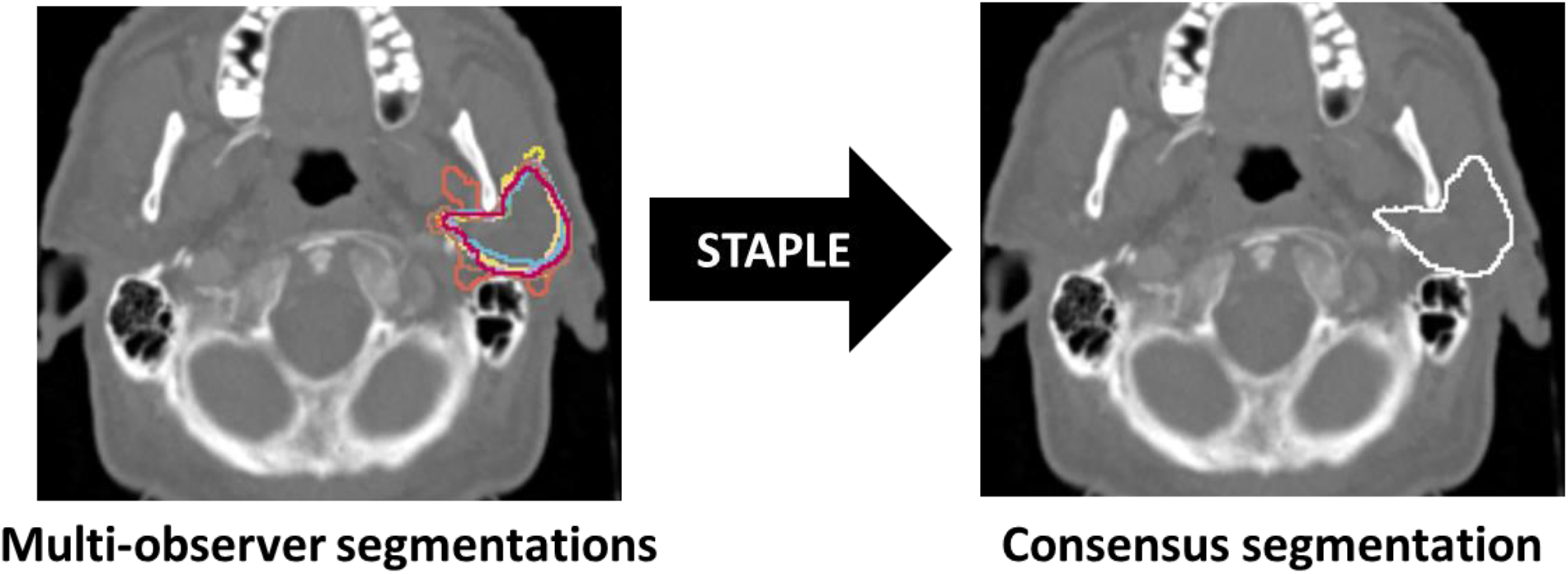
Expert segmentations of the left parotid gland in the head and neck case (left) and corresponding simultaneous truth and performance level estimation (STAPLE) consensus segmentation (right).

### Similarity Metric Computations

To compare ROI segmentation quality, we implemented various metrics of geometric similarity. For our analysis, we focused on the Dice similarity coefficient (DSC), a well-established volume-based metric for segmentation studies, and the surface DSC (SDSC), a newer surface distance metric that has been shown to be germane to potentially improving radiation oncology workflows, particularly for time savings [25,26]. Metrics were calculated using the surface-distances Python package [27] and in-house Python code. SDSC was calculated based on ROI-specific thresholds determined by measuring the median pairwise mean surface distance of all expert segmentations for that ROI as suggested in literature [27]; tolerance values, required parameters for SDSC calculation, used for each ROI are shown in **Appendix C**. Additional segmentation similarity metrics, including the 95% Hausdorff distance and added path length were also investigated in supplementary analyses (**Appendix D**). Pairwise metric calculations within a group (non-expert, expert) were used to determine interobserver metric values for DSC and SDSC. The median interobserver value for experts was considered as a theoretical threshold of clinical acceptability. Metric values were also computed between the expert STAPLE segmentations and the non-expert STAPLE segmentations. ROI volumetric comparisons between non-experts and experts were also investigated in **Appendix E**.

### Non-Expert STAPLE bootstrap experiments

To determine the number of non-experts required to cross the mean interobserver metric value, we performed a bootstrap resampling experiment where random subsets of non-experts were selected with replacement to generate a STAPLE consensus segmentation and compared against the expert STAPLE segmentation. Experiments were conducted for 2, 3, 4, 5, 6, 7, 8, 9, 10, 15, 20, and 25 non-expert subsets. 100 bootstrap iterations were conducted for each ROI to construct 95% confidence intervals. Bootstrap iterations took between 10-12 hours for each ROI on a standard central processing unit (Intel® Core™ i7-8700 Processor). Bootstrap results were displayed as line plots.

### Statistical Analysis

Pairwise metrics were compared between non-experts and experts using Mann Whitney U tests using the Python statannotations package; Mann Whitney U tests were selected due to the non-normal distribution of data and imbalance of sample sizes between experts and non-experts [28].

### Code and Data Sharing

All analysis code is available online in the form of Jupyter Notebooks through GitHub repositories: https://github.com/kwahid/C3RO_analysis (private until manuscript acceptance). Anonymised data used in our analysis are made publicly available on Figshare, doi: 10.6084/m9.figshare.21074182 (private until manuscript acceptance).

## Results

As of August 2022, C3RO had 1,026 unique registrants, 221 of whom contoured at least one case. Among the participants who contoured, 127 (57%) identify as male and 93 (42%) identify as female. Participant race and ethnic backgrounds are as follows: 96 (43%) White; 80 (36%) Asian or Pacific Islander; and 22 (10%) Hispanic, Latino, or Spanish origin; 7 (3%) Black. Only 52 (24%) of participants are from the United States, while 169 (76%) of the participants are international. Most, 169 (76%), participants are practicing radiation oncologists, while 40 (18%) are resident physicians, 7 (3%) are radiation therapists, and 1 (<1%) is a medical physicist. The median (IQR) years of experience is 5 (3, 10) for attending physicians after residency and is 3 (2, 4) for resident physician year in residency. Most, 146 (66%), participants work in an academic setting or are affiliated with a University, 50 (23%) work in a non-academic hospital, and 21 (10%) are part of private practice. Participant characteristics can be found in **Table 2**.

**Table 2.** Participant Characteristics.

|  | Case 1:<br>Breast | Case 2:<br>Sarcoma | Case 3:<br>H&N | Case 4:<br>GYN | Case 5:<br>GI | Participants who<br>contoured | All Registrants |
| --- | --- | --- | --- | --- | --- | --- | --- |
| Number of participants, n | 132 | 66 | 81 | 49 | 30 | 221 | 1026 |
| Gender, n (%) |  |  |  |  |  |  |  |
| • Male | 78 (59%) | 39 (59%) | 42 (52%) | 29 (59%) | 14 (47%) | 127 (57%) | 546 (53%) |
| • Female | 54 (41%) | 27 (41%) | 38 (47%) | 20 (41%) | 16 (53%) | 93 (42%) | 465 (45%) |
| • Other | 0 (0%) | 0 (0%) | 1 (1%) | 0 (0%) | 0 (0%) | 1 (<1%) | 19 (2%) |
| Race/Ethnicity*, n |  |  |  |  |  |  |  |
| • White | 58 (44%) | 30 (45%) | 37 (46%) | 20 (41%) | 15 (50%) | 96 (43%) | 385 (38%) |
| • Asian or Pacific Islander | 46 (35%) | 23 (35%) | 25 (31%) | 15 (31%) | 10 (33%) | 80 (36%) | 347 (34%) |
| • Hispanic, Latino, or Spanish origin | 13 (10%) | 6 (9%) | 8 (10%) | 8 (16%) | 3 (10%) | 22 (10%) | 123 (12%) |
| • Black | 5 (4%) | 1 (2%) | 3 (4%) | 2 (4%) | 3 (10%) | 7 (3%) | 55 (5%) |
| • Other | 12 (9%) | 7 (11%) | 10 (12%) | 4 (8%) | 3 (10%) | 21 (10%) | 148 (14%) |
| Geographic setting, n (%) |  |  |  |  |  |  |  |
| • US | 30 (23%) | 14 (21%) | 14 (17%) | 8 (16%) | 6 (20%) | 52 (24%) | 197 (19%) |
| • Non-US | 102 (77%) | 52 (79%) | 67 (83%) | 41 (84%) | 24 (80%) | 169 (76%) | 829 (81%) |
| Profession, n (%) |  |  |  |  |  |  |  |
| • Radiation Oncologist / Clinical Oncologist | 102 (77%) | 47 (71%) | 64 (79%) | 37 (76%) | 21 (70%) | 169 (76%) | 740 (72%) |
| • Resident Physician | 24 (18%) | 15 (23%) | 12 (15%) | 7 (14%) | 5 (17%) | 40 (18%) | 210 (20%) |
| • Radiation Therapist | 3 (2%) | 2 (3%) | 2 (2%) | 2 (4%) | 2 (7%) | 7 (3%) | 33 (3%) |
| • Medical Student | 0 (0%) | 0 (0%) | 0 (0%) | 0 (0%) | 0 (0%) | 0 (0%) | 6 (<1%) |
| • Medical Physicist | 0 (0%) | 0 (0%) | 1 (1%) | 0 (0%) | 0 (0%) | 1 (<1%) | 9 (<1%) |
| • Other | 3 (2%) | 2 (3%) | 2 (2%) | 3 (6%) | 2 (7%) | 4 (2%) | 28 (3%) |
| Years of experience, median (IQR) |  |  |  |  |  |  |  |
| • Attending physician years after residency | 4 (2, 8) | 3 (1.75, 8) | 6 (2, 11.75) | 6 (3, 11) | 7 (4, 14) | 5 (3, 10) | 5 (3, 10) |
| • Resident physician year in residency | 3 (2, 4) | 2 (1, 3) | 2.5 (2, 3.25) | 2 (2, 3) | 3 (3, 4) | 3 (2, 4) | 3 (2, 4) |
| Practice type, n (%) |  |  |  |  |  |  |  |
| • Academic / University | 83 (63%) | 40 (61%) | 50 (62%) | 32 (65%) | 16 (53%) | 146 (66%) | 651 (63%) |
| • Non-academic Hospital | 35 (27%) | 15 (23%) | 22 (27%) | 10 (20%) | 5 (17%) | 50 (23%) | 219 (21%) |
| • Private Practice (solo or group) | 13 (10%) | 8 (12%) | 8 (10%) | 6 (12%) | 6 (20%) | 21 (10%) | 121 (12%) |

\* indicates that participants could select multiple options

### Interobserver variability

Interobserver variability of non-experts and experts based on pairwise segmentation comparisons are shown in **Figure 3**. For the breast case, the interobserver DSC of 3/7 ROIs (CTV_Ax, CTV_IMN, and CTV_Sclav_LN) and interobserver SDSC of 2/7 ROIs (CTV_Ax and CTV_IMN) were significantly higher for experts vs. non-experts. For the sarcoma case, the interobserver DSC and SDSC of 1/3 ROIs (GTV) was significantly higher for experts vs. non-experts. For the H&N case, the interobserver DSC and SDSC of 10/11 ROIs (GTVp, GTVn, CTV1, CTV2, Brainstem, Glnd_Submand_L, Glnd_Submand_R, Musc_Constrict, parotid_l, parotid_r) were significantly higher for experts vs. non-experts. For the GYN case, the interobserver DSC and SDSC of 2/4 ROIs (CTVn_4500, Bowel_Small) were significantly higher for experts vs. non-experts. For the GI case, the interobserver DSC of 2/3 ROIs (CTV_4500, CTV_5400) was significantly higher for experts vs. non-experts. The remaining ROIs for all cases were non-significantly different between experts and non-experts. Volumetric comparisons for ROIs between non-experts and experts are shown in **Appendix E**; only 3 ROIs were significantly different between non-experts and experts, namely, the breast case CTV_IMN, the sarcoma case GTV, and the H&N case Parotid_L.

**Figure 3.**
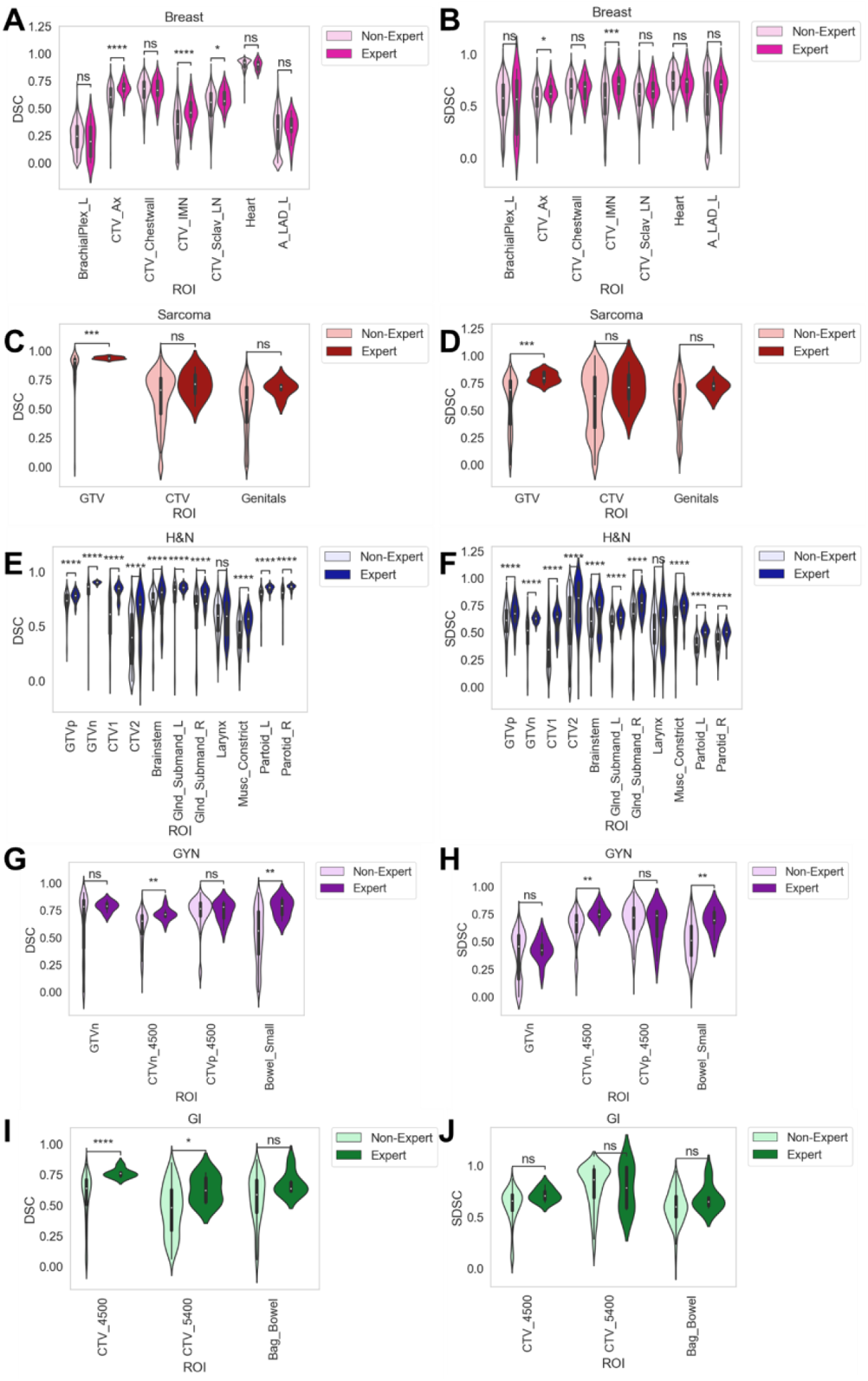
Interobserver variability based on pairwise segmentation comparisons for radiation oncologists of varying expertise (non-expert, expert). Breast, sarcoma, H&N, GYN, and GI cases are shown in the 1st, 2nd, 3rd, 4th, and 5th rows, respectively. DSC and surface DSC metrics shown in left and right panels, respectively. Stars above plot indicate Mann Whitney U test level of significance: ns: p > 0.05; *: 0.01 < p <= 0.05; **: 0.001 < p <= 0.01; ***: 0.0001 < p <= 0.001; ****: p <= 0.0001. Abbreviations: H&N = head and neck, GYN = gynecology, _l = left, _r = right, gtv = gross tumor volume, ctv = clinical target volume.

### STAPLE comparisons

Comparisons of consensus segmentations for all non-expert segmentations vs. consensus segmentations for all expert segmentations are shown in **Figure 4**. For the breast case, non-expert consensus segmentations for all 7 ROIs crossed the expert interobserver values for both DSC and SDSC. For the sarcoma case, non-expert consensus segmentations for 2 out of 3 ROIs (GTV, CTV) crossed the expert interobserver values for both DSC and SDSC. For the H&N case, non-expert consensus segmentations for 9 out of 11 ROIs (GTVp, GTVn, CTV2, Glnd_Submand_L, Glnd_Submand_R, Larynx, Musc_Constrict, parotid_l, parotid_r) crossed the expert interobserver DSC, while 7 out of 11 ROIs (GTVn, CTV2, Glnd_Submand_L, Larynx, Musc_Constrict, parotid_l, parotid_r) crossed the expert interobserver SDSC. For the GYN case, non-expert consensus segmentations for all 4 ROIs crossed the expert interobserver values for both DSC and SDSC. For the GI case, non-expert consensus segmentations for 2 out of 3 ROIs (CTV_4500, Bag_Bowel) crossed the interobserver DSC, while 2 out of 3 ROIs (CTV_4500, CTV_5400) crossed the expert interobserver SDSC.

**Figure 4.**
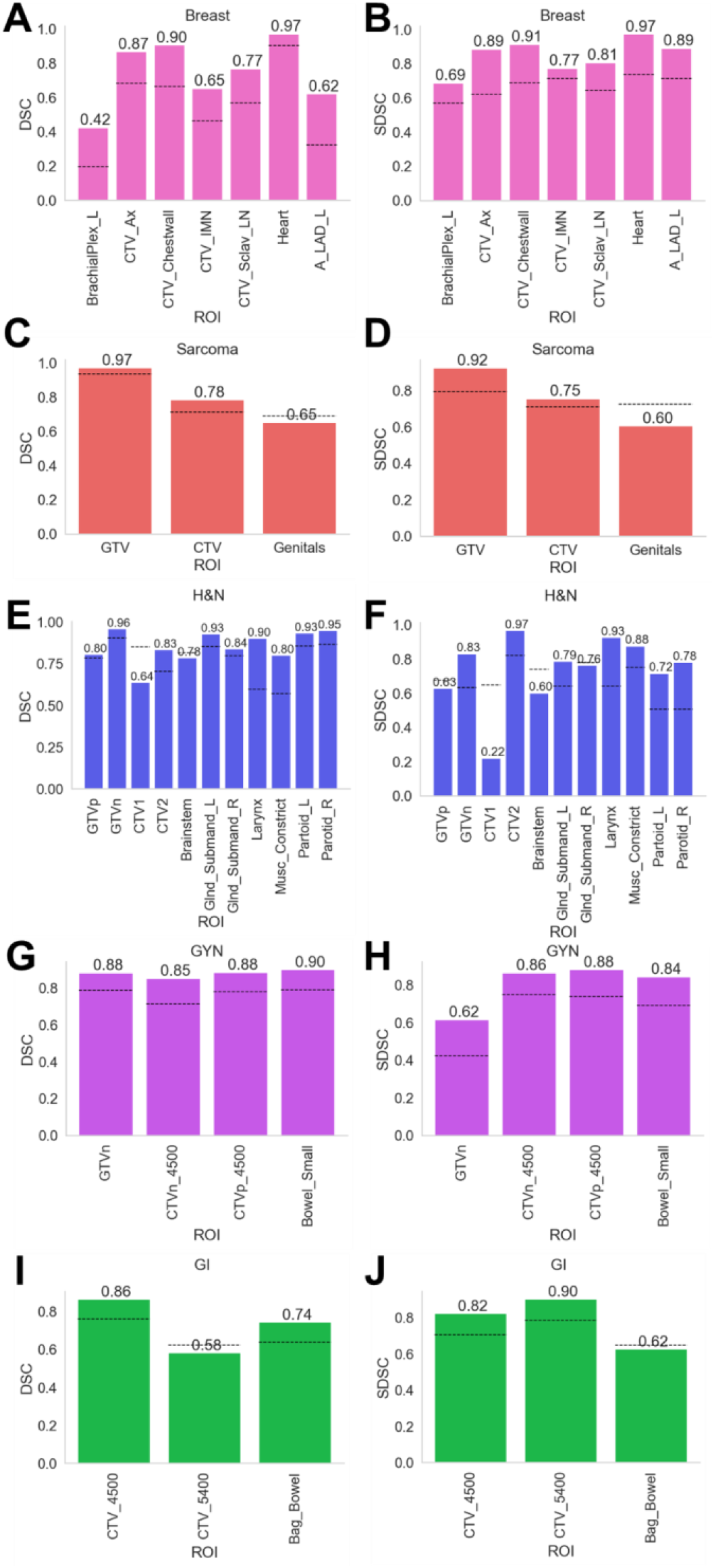
Dice similarity coefficient (DSC) and surface DSC (SDSC) values comparing non-expert STAPLE using maximum number of available cases to expert STAPLE. Breast, sarcoma, H&N, GYN, and GI cases are shown in the 1st, 2nd, 3rd, 4th, and 5th rows, respectively. DSC and SDSC metrics shown in left and right panels, respectively. Black dotted lines indicate median expert interobserver value for that metric. Abbreviations: H&N = head and neck, GYN = gynecology, _l = left, _r = right, gtv = gross tumor volume, ctv = clinical target volume.

### STAPLE visual comparisons

We visually investigated 1 ROI in the H&N case which exhibited outlier behavior, namely CTV1. For both DSC and SDSC, the non-expert STAPLE of CTV1 was unable to cross the corresponding expert interobserver values. As shown in **Figure 5**, the expert STAPLE generally led to a more conservative estimate of the ROI, compared to the non-expert STAPLE which covered a greater area. For completeness, we also show CTV2 for both experts and non-experts, which also showed more conservative estimates for experts vs. non-experts.

**Figure 5.**
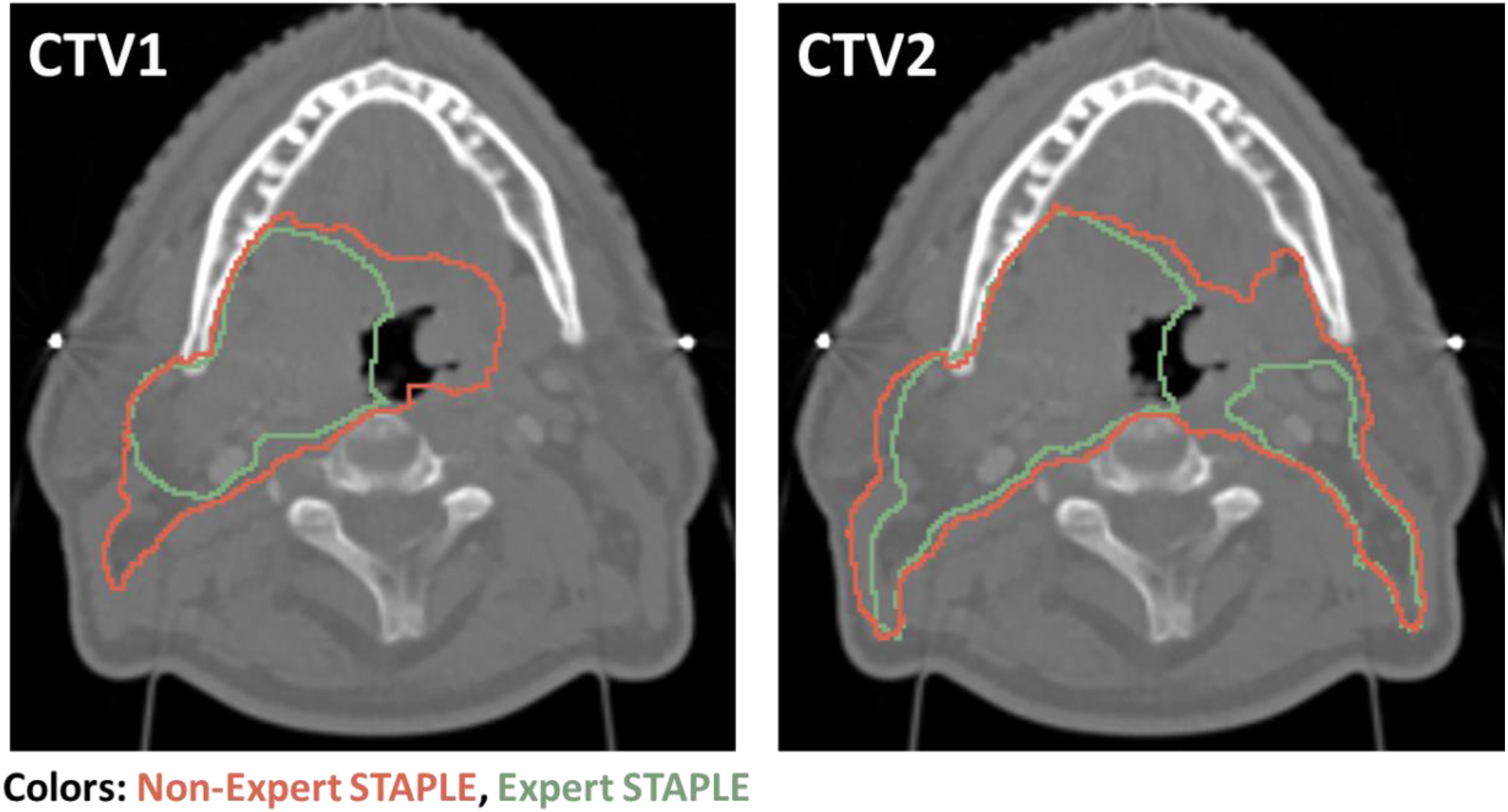
Expert STAPLE (green) and non-expert STAPLE (red) for CTV1 (left) and CTV2 (right) for head and neck case.

### Non-expert STAPLE bootstrap Experiments

Non-expert STAPLE bootstrap experiments for the breast, sarcoma, H&N, GYN, and GI cases are shown in **Figure 6**. For the breast case, expert interobserver DSC was crossed between a minimum 2-4 observers across the various ROIs; the smallest minimum number of observers (2) was achieved for BrachialPlex_L and Heart, while the largest minimum number of observers (4) was achieved for CTV_Ax and CTV_IMN. For the sarcoma case, expert interobserver DSC was crossed between a minimum 3-5 observers across the various ROIs; the smallest minimum number of observers (3) was achieved for genitals, while the largest minimum number of observers (5) was achieved for CTV. For the H&N case, expert interobserver DSC was crossed between a minimum 3-5 observers across the various ROIs; the smallest minimum number of observers (3) was achieved for GTVn, Brainstem, Glnd_Submand_L, Glnd_Submand_R, and Larynx, while the largest minimum number of observers (5) was achieved for musc_constric_all and parotid_l. For the GYN case, expert interobserver DSC was crossed between a minimum 3-5 observers across the various ROIs; the smallest minimum number of observers (3) was achieved for GTVn, while the largest minimum number of observers (5) was achieved for CTVn_4500. For the GI case, expert interobserver DSC was crossed using a minimum 3 observers for all ROIs. Heatmap representations of bootstrap experiments can be found in **Appendix D, Figure D18**. Of note, the following ROIs showed non-saturating performance with an increasing number of non-experts used in the consensus segmentation: breast (BrachialPlex_L: highest non-expert DSC = 0.63, non-expert DSC using all observers = 0.42), sarcoma (genitals: highest non-expert DSC = 0.83, non-expert DSC using all observers = 0.60), H&N (GTVp: highest non-expert DSC = 0.87, non-expert DSC using all observers = 0.80; CTV1: highest non-expert DSC = 0.86, non-expert DSC using all observers = 0.64), GYN (GTVn: highest non-expert DSC = 0.90, non-expert DSC using all observers = 0.88), GI (CTV_5400: highest non-expert DSC = 0.78, non-expert DSC using all observers = 0.58; Bag_Bowel: highest non-expert DSC = 0.85, non-expert DSC using all observers = 0.74). Bootstrap experiments for additional metrics in line plot format can be found in **Appendix D**.

**Figure 6.**
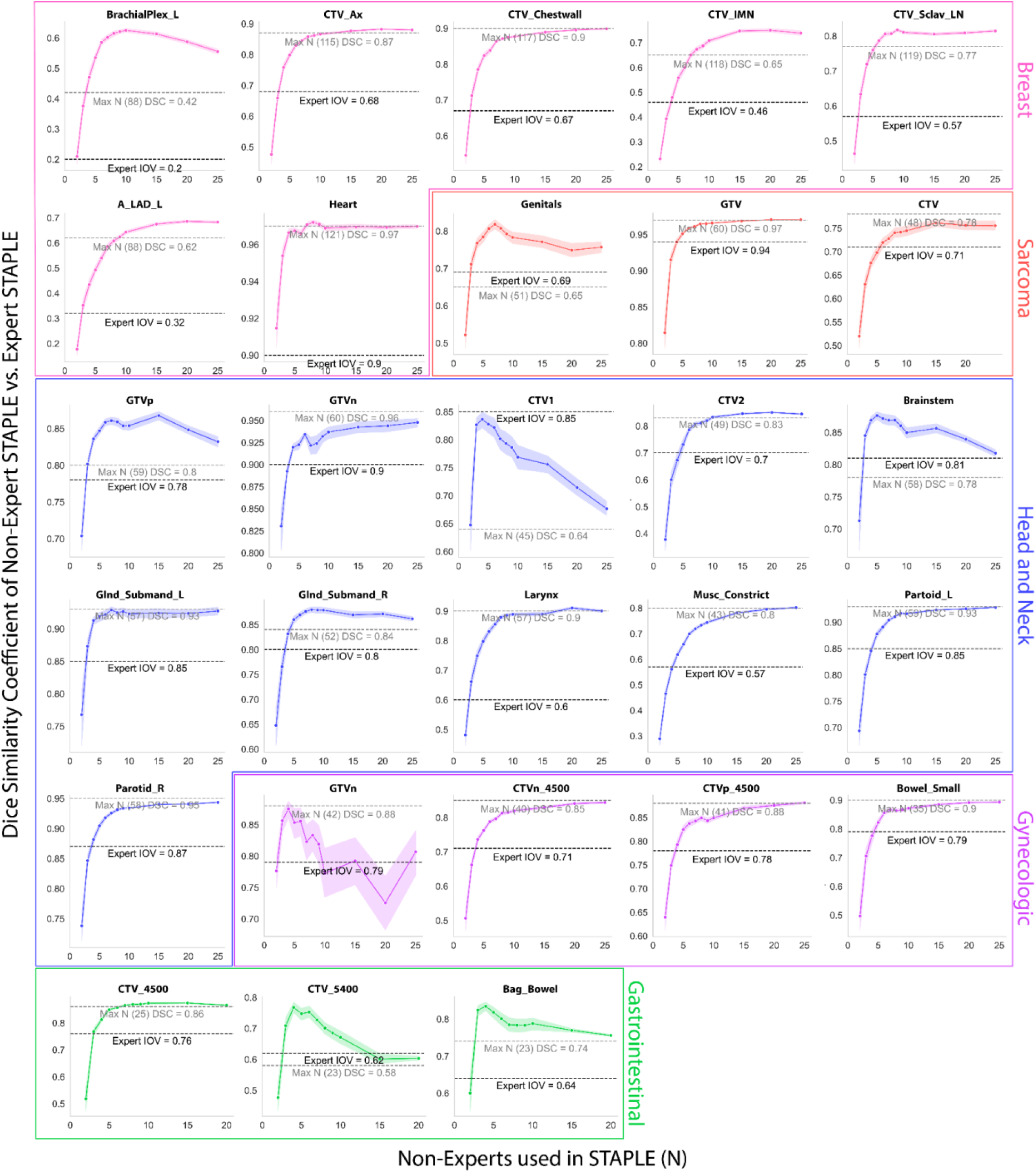
Consensus segmentation bootstrap experiments. Pink, red, blue, purple, and green plots correspond to breast, sarcoma, head and neck, gynecologic, and gastrointestinal regions of interest, respectively. Black dotted lines indicate median expert interobserver dice similarity coefficient (DSC) for a corresponding region of interest. Gray dotted lines indicate DSC performance using the maximum number of non-experts in the consensus segmentation.

## Discussion

In this study, we have systematically investigated the difference between non-experts and experts in contouring ROIs for several disease sites using various evaluation metrics. We have probed the inherent interobserver variability within non-experts and experts and determined several ROIs have better agreement when contoured by experts. Consensus segmentation experiments reveal that most consensus expert ROI contours can be roughly approximated using non-expert segmentations, which cross expert interobserver group variability performance thresholds. Our results provide justification towards using large-scale non-expert contours for “gold-standard” segmentation data in the absence of multiple expert “ground-truth” data availability, and glean insight into the behavior of consensus contours across a large number of observer inputs. While crowdsourcing is common in medical image analysis [29], there have been few studies evaluating the use of crowdsourcing for contour quality. To our knowledge, this is the largest study characterizing segmentation performance across multiple physician observers, and the first study to investigate crowdsourced contour performance in the context of radiation oncology workflows.

Our interobserver variability experiments demonstrate that several ROIs across the various cases have higher interobserver agreement for experts compared to those of non-experts for both volumetric and surface distance metrics. Generally, the interobserver variability did not vary significantly among the OAR structures for most cases. Key outliers to these trends were the majority of H&N case OAR ROIs and the Bowel_Small ROI of the GYN case. Analogously, target volumes tended to be among the ROIs that were significantly different between non-expert and expert interobserver variability. It is well known that tumor-related tissues are inherently more heterogeneous than healthy tissues. As such, our results may be explained by the potential higher subjectivity of target volume contours compared to those of OARs. A previous study by Cardenas et al. investigating large-scale multi-observer segmentation in H&N cancer using magnetic resonance imaging supports our results, whereby target volumes demonstrated particularly low agreement between observers compared to OARs [30]. Nonetheless, it warrants highlighting that certain cases exhibited a greater predilection for improved contour consistency in experts compared to those of non-experts. This is most apparent in the H&N case, where the vast majority of ROIs (10/11) showed higher agreement for experts compared to those of non-experts. These findings are congruent with previous literature indicating that H&N is a particularly challenging disease site for physician-based contouring [9,19,31].

Our initial investigations comparing STAPLE consensus segmentations using all non-expert observers against the STAPLE consensus segmentation using all expert observers revealed that non-expert STAPLE contours could cross expert interobserver variability for most ROIs; generally, there was strong agreement between results for volumetric and surface distance metrics. However, a few key outliers were unable to cross interobserver variability, namely, the genitals ROI (DSC and SDSC) for the sarcoma case, the GTVp (SDSC only), CTV1 (DSC and SDSC), Brainstem (SDSC only), and Glnd_Submand_L (SDSC only) ROIs for the H&N case, and the CTV_5400 (DSC only) and Bag_Bowel (SDSC only) ROIs for the GI case. Interestingly, unlike the interobserver variability analysis, where target volumes tended to have greater variability among non-experts, this trend did not necessarily translate when considering consensus segmentations of all the non-expert observers. Importantly, one particular ROI that had a large degree of difference between non-expert and expert STAPLE consensus contours was CTV1 of the H&N case. Upon visual investigation, the difference between non-expert and expert STAPLE segmentations was likely due to considerable non-expert uncertainty stemming from the inclusion of two individual subregions (GTVn and GTVp) in addition to areas of microscopic tumor spread, thereby leading to a larger consensus segmentation for the non-experts. These differences became less apparent when asking non-experts to contour CTV2, as the incorporation of non-ambiguous tissue (lymph node levels) seemingly increased the conformity between observers. It is worthy to note that in Cardenas et al. individual differences between GTV and CTV ROIs were minimal and are likely attributed to separate CTVs generated for primary and nodal tumors [30], as opposed to our study where a single CTV was generated combining both primary and nodal volumes.

Having confirmed that non-expert consensus segmentations could approximate expert consensus segmentations to a reasonable degree by crossing expert interobserver variability cutoffs, we sought to determine how the dynamics of segmentation performance were affected by the number of non-experts used in the STAPLE algorithm. For most ROIs, there was a general trend that on the order of 2-5 non-experts were needed to cross expert interobserver variability. Our results are congruent with previous literature on crowdsourcing labels for pathological patterns in lung imaging, where a limited number of observers could be combined through consensus methods to match reference repeatability [32]. Consistent with trends observed in our interobserver variability experiments, the ROIs that required the greatest number of non-experts to cross expert interobserver variability cutoffs were often target volumes, while the ROIs that required the least number of non-experts were OARs. The majority of ROIs exhibited a maximum DSC value, i.e., performance saturation, at a certain optimal number of non-experts used in the STAPLE algorithm, which then plateaued and maintained this high performance up to the maximum number of observers used. However, a small number of ROIs exhibited non-saturating performance effects, where after maximum performance was achieved the addition of a greater number of observers in the STAPLE algorithm decreased performance, often precipitously. As before, these tended to be target volumes where a large degree of heterogeneity between the non-experts was expected, but also included a few OARs that would be considered particularly challenging because of heterogeneity in visual interpretation, e.g., brachial plexus (breast case) and genitals (sarcoma case).

In the medical image segmentation space, several studies have been conducted on the use of “noisy” labels for model training, with mixed results. Within radiation oncology, one study in particular has shown that for at least OAR contours, deep learning may be robust to noisy segmentations [33]. A similar study investigating cardiac segmentation on ultrasound found that the training of deep learning models with novice data was not significantly different from deep learning models trained with expert data [34]. Additionally, the authors show average DSC for expert-expert and novice-expert segmentations show that no statistical difference is found between the variability of annotators in several of the annotations of interest, results which are echoed in a recent study investigating crowdsourcing for liver tumor segmentation where the quality of annotations was not statistically significantly different four distinct groups [35]. Contrary to the previous studies, Wesemeyer et al. demonstrate a tradeoff between quality and quantity for deep learning segmentation performance [36]. As we demonstrate in our study, expert consistency may be better than non-expert consistency for some radiotherapy-related ROIs, particularly for H&N imaging. Therefore, there is still likely a need to generate “expert-level” gold-standard contours in training radiotherapy-related deep learning models, at least for select cases or structures. Our study demonstrates that the use of consensus contours using non-experts may be a reasonable approximation to gold-standard expert contours using a relatively small number of observers, and may exhibit particular utility in scenarios where auto-segmentation models require large amounts of data but only a limited number of experts are available to provide segmentations.

Our study has several limitations. Firstly, we have only investigated one case per disease site in a single imaging modality (computed tomography). Therefore, our results may not necessarily generalize to arbitrary cases in different imaging modalities (e.g., magnetic resonance imaging, positron emission tomography, etc.), particularly for target volumes which can exhibit significant heterogeneity between cases and modalities. However, given that computed tomography is the current gold-standard for radiotherapy planning, we believe our results are of significant interest to the radiation oncology community. There was no contouring protocol provided to the participants for reference in this challenge, which may have reduced optimal performance. To evaluate contours, we utilized geometric indices, which are not well correlated with clinically meaningful endpoints; rather, a multi-domain approach including dosimetric indices and clinical input has been shown to be the best method to evaluate auto-segmentation [25]. An additional limitation of our study is that we have stratified physician expertise based on subjective criteria. Herein, we have defined an expert as an individual who is recognized in their field and/or contributed to consensus guideline generation. While there may be additional methodology to stratify expertise, we have chosen this method as it separates physicians based on perceived familiarity with standardized guidelines. Nonetheless, despite this limitation, we show a difference between this stratification of experts and non-experts. More studies are needed to determine objective criteria for expertise. We fully acknowledge this may mischaracterize some individuals into “non-expert” categories erroneously, even if they have significant familiarity with guidelines, but plan to investigate alternative stratifications based on additional objective criteria in future studies. Moreover, our current analysis only investigates radiation oncologists who have at some level completed formal training. We have not investigated novice observers, where observers may have no formal education in medical anatomy or segmentation. Future studies should investigate novice observers as these labels would potentially be the most cost effective to obtain and may also be able to approximate the current gold-standard.

Importantly, our study provides a large high-quality curated dataset that can act as a reference for future studies on interobserver contour variability and auto-segmentation in radiation oncology workflows. Moreover, we publicly distribute our raw imaging data and open-source our analysis pipelines so the community can investigate these claims further. Finally, our results highlight the differences in non-expert and expert contours, which can be further leveraged to create educational tools for trainee segmentation quality control.

## Conclusions

In summary, using five distinct disease sites (breast, sarcoma, H&N, GYN, and GI), we have systematically investigated differences in contour quality between non-expert and expert radiation oncologist observers in target volumes and OARs. Overall, there was a general trend towards experts providing more consistent segmentations in terms of pairwise DSC and SDSC for a variety of ROIs. Moreover, we showed that using the STAPLE algorithm, consensus contours could be generated from non-experts that approximate gold-standard expert segmentations to a reasonable degree (crossing expert interobserver variability) for most ROIs; some target volumes were unable to be approximated readily. Finally, we experimented with a variable number of non-experts in generating consensus contours and demonstrated for most ROIs 2-5 non-experts is sufficient to cross expert interobserver variability, though specific attention should be paid to ROIs such as target volumes and complex OARs which exhibit decreased performance as more observers are added to the consensus segmentation. Our study acts as a potential reference for the characterization of interobserver variability and use of consensus contours in future AI-related radiotherapy applications. Future work will include the investigation of a greater number of disease sites, cases, imaging modalities, and levels of expertise.

## Supporting information

Other Appendices

## Data Availability

Anonymized data used in our analysis are made publicly available on Figshare, doi: 10.6084/m9.figshare.21074182 (private until manuscript acceptance).

