## Appendix A for "“E Pluribus Unum”: Prospective acceptability benchmarking from the Contouring Collaborative for Consensus in Radiation Oncology (C3RO) Crowdsourced Initiative for Multi-Observer Segmentation"

### C3RO Intake Survey

Resize font:

Page 1 of 1

**First name**

**Last name**

**Hospital/organization**

**Country**

**Email address (for gift card raffle)**

**Why are you interested in participating in this challenge?**
*Select all that apply*

- ☐ Opportunity to win gift cards
- ☐ Comparison to peers/leaderboard
- ☐ Access to educational content
- ☐ Access to contour datasets for guideline/research purposes

**How did you first hear about this challenge?**
*Select all that apply*

- ◯ Twitter
- ◯ eContour website announcement
- ◯ Email from eContour or ProKnow
- ◯ Colleague (word of mouth)
- ◯ ROECSG newsletter
- ◯ ROECSG annual symposium
- ◯ Other

reset

**What is your profession?**

- ◯ Radiation Oncologist/Clinical Oncologist
- ◯ Resident Physician
- ◯ Medical Student
- ◯ Medical Physicist
- ◯ Dosimetrist
- ◯ Radiation Therapist
- ◯ Other

reset

**What year did you start practicing (graduate residency)?**

**Gender**

**Which categories describe you?**
*Select all that apply*

- ☐ White
- ☐ Hispanic, Latino, or Spanish origin
- ☐ Black or African American
- ☐ Asian
- ☐ Other

**Do you have an academic affiliation?**

- ◉ Yes
- ◯ No

reset

**Which best describes your primary practice?**

- ◯ Academic/university
- ◯ Non-academic hospital
- ◯ Private practice (solo or group)
- ◯ Other

reset

**How many radiation oncologist colleagues do you work with at your primary site (excluding you)?**

[ ]

**On most days you are in clinic, is there another radiation oncologist on site with you?**

◉ Yes ◯ No

reset

**Which disease site(s) do you treat?**
*Select all that apply*

- [ ] Breast
- [ ] CNS
- [ ] Gastrointestinal
- [x] Genitourinary
- [ ] Gynecologic
- [ ] Head & Neck
- [ ] Lung, Thoracic
- [ ] Lymphoma, Leukemia
- [ ] Metastatic
- [ ] Pediatric
- [ ] Sarcoma
- [ ] Skin, Cutaneous

**Approximately how many patients with genitourinary cancer do you treat per month?**

[ ]

**Do you consider yourself an "expert" in any of the aforementioned disease sites?**

- ◉ Yes
- ◯ No

reset

**Please list area(s) and years of experience with that area**

[ ]

Expand

**By submitting this survey, you consent to the use of the de-identified results and responses you provide in aggregate with other participants' results and responses for analysis. Participation in this challenge is voluntary and at no cost to you.**

◯ I acknowledge and accept these terms

reset

Submit

Save & Return Later
