## Supplementary material for "“E Pluribus Unum”: Prospective acceptability benchmarking from the Contouring Collaborative for Consensus in Radiation Oncology (C3RO) Crowdsourced Initiative for Multi-Observer Segmentation": Other Appendices

### Appendix B. Imaging Characteristics

**Table B1.** Computed tomography (CT) acquisition parameters for C3RO cases.

| CT Acquisition Parameter | Breast | Sarcoma | H&N | GYN | GI |
|---|---|---|---|---|---|
| Manufacturer | GE | SIEMENS | SIEMENS | GE | GE |
| Model | Discovery CT590 RT | SOMATOM Confidence | Sensation Open | Discovery CT590 RT | Discovery CT590 RT |
| Slice Thickness (mm) | 2.5 | 3 | 3 | 2.5 | 2.5 |
| KVP (kV) | 120 | 120 | 120 | 120 | 120 |
| Exposure Time (ms) | 891 | 1000 | 1000 | 856 | 856 |
| X-Ray Tube Current (mA) | 154 | 111 | 32 | 167 | 277 |
| Rows | 512 | 512 | 512 | 512 | 512 |
| Columns | 512 | 512 | 512 | 512 | 512 |
| In-plane Resolution (mm) | 1.26 | 1.26 | 0.98 | 0.98 | 0.98 |
| Reconstruction diameter (mm) | 650 | 650 | 500 | 500 | 500 |
| Number of axial slices | 140 | 229 | 143 | 195 | 196 |

### Appendix C. Tolerance Values

**Table C1.** Tolerance values used for surface Dice similarity coefficient value computations for each region of interest (ROI).

| Case | ROI | Tolerance Value (mm) |
|---|---|---|
| Breast | BrachialPlex_L | 4.2 |
| | CTV_Ax | 3.4 |
| | CTV_Chestwall | 4.4 |
| | CTV_IMN | 2.7 |
| | CTV_Sclav_LN | 3.6 |
| | Heart | 1.8 |
| | A_LAD_L | 4.6 |
| Sarcoma | GTV | 0.4 |
| | CTV | 5.4 |
| | Genitals | 3.2 |
| H&N | GTVp | 1.4 |
| | GTVn | 0.5 |
| | CTV1 | 1.3 |
| | CTV2 | 10.3 |
| | Brainstem | 1.1 |
| | Glnd_Submand_L | 0.5 |

| | | |
|---|---|---|
| | Glnd_Submand_R | 1.0 |
| | Larynx | 2.2 |
| | Musc_Constrict | 2.1 |
| | Partoid_L | 0.9 |
| | Parotid_R | 0.8 |
| GYN | GTVn | 0.9 |
| | CTVn_4500 | 3.0 |
| | CTVp_4500 | 3.8 |
| | Bowel_Small | 5.0 |
| GI | Bag_Bowel | 6.12 |
| | CTV_4500 | 4.09 |
| | CTV_5400 | 15.22 |

### Appendix D. Additional Metric Representations

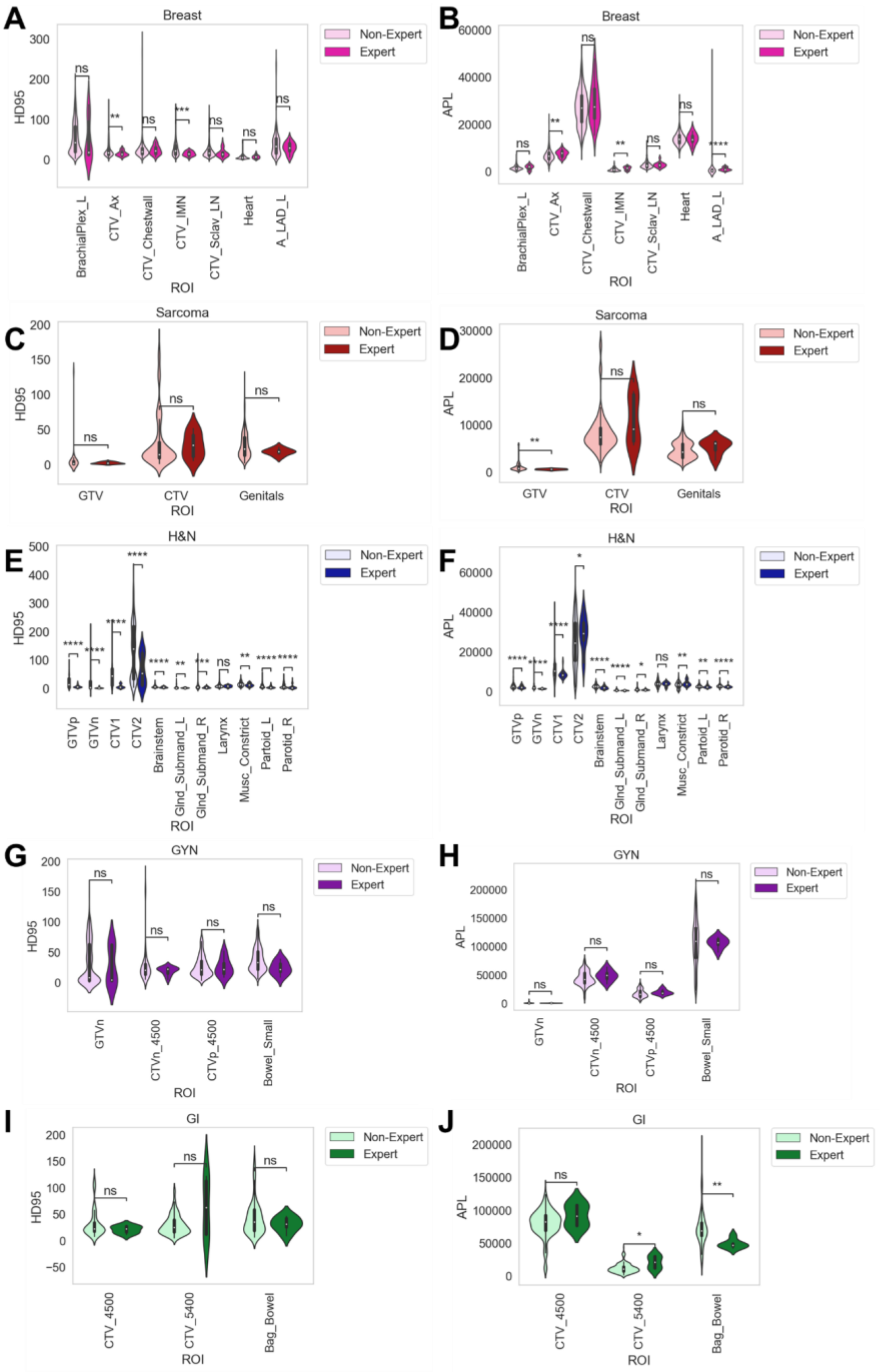


**Figure D1.** Additional interobserver variability plots for 95% Hausdorff distance (HD95) and added path length (APL).

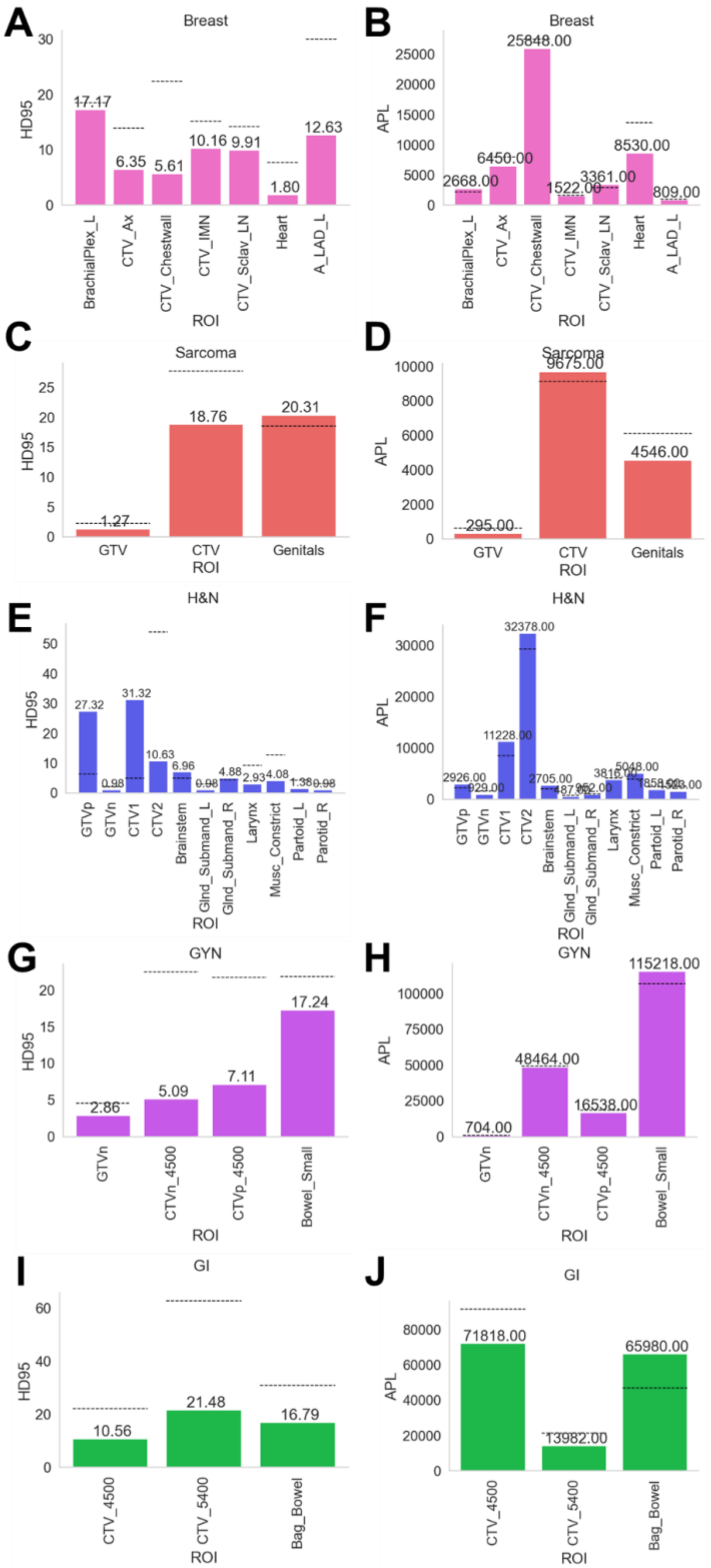


**Figure D2.** Additional metrics comparing non-expert STAPLE using maximum number of available cases to expert STAPLE. 95% Hausdorff distance (HD95) and added path length (APL).

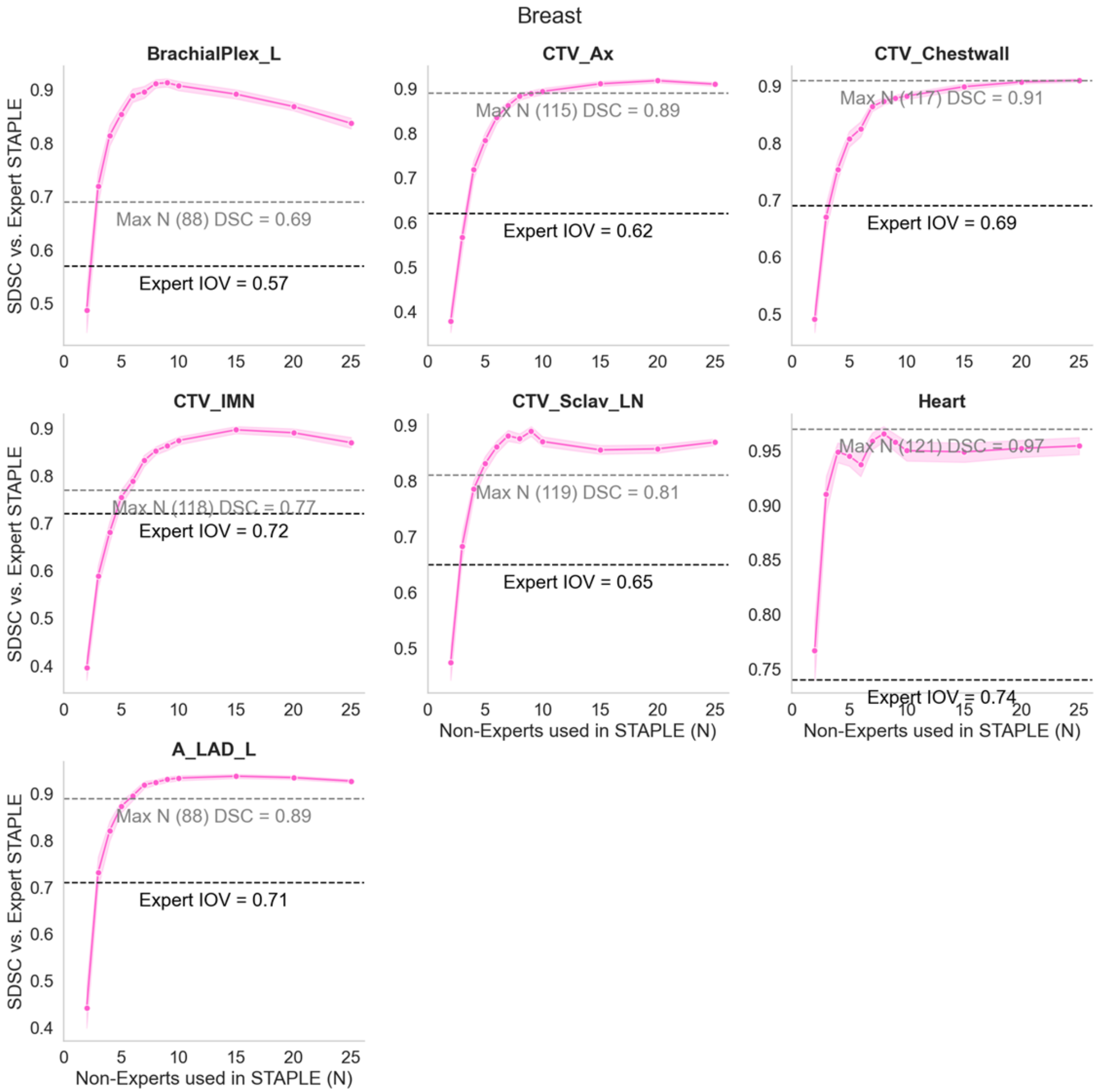


**Figure D3.** Bootstrap experiment results for Breast case using SDSC.

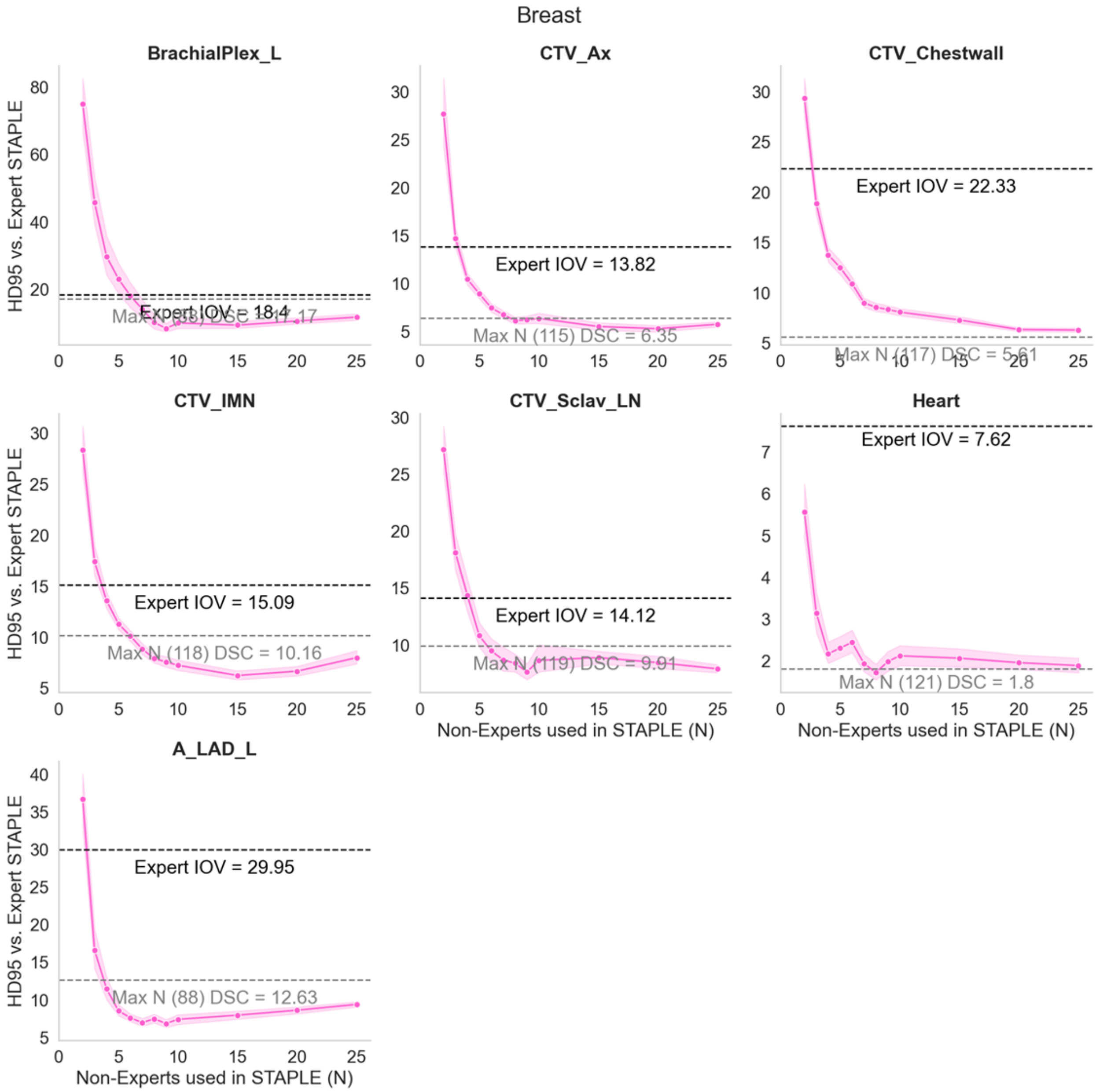


**Figure D4.** Bootstrap experiment results for Breast case using HD95.

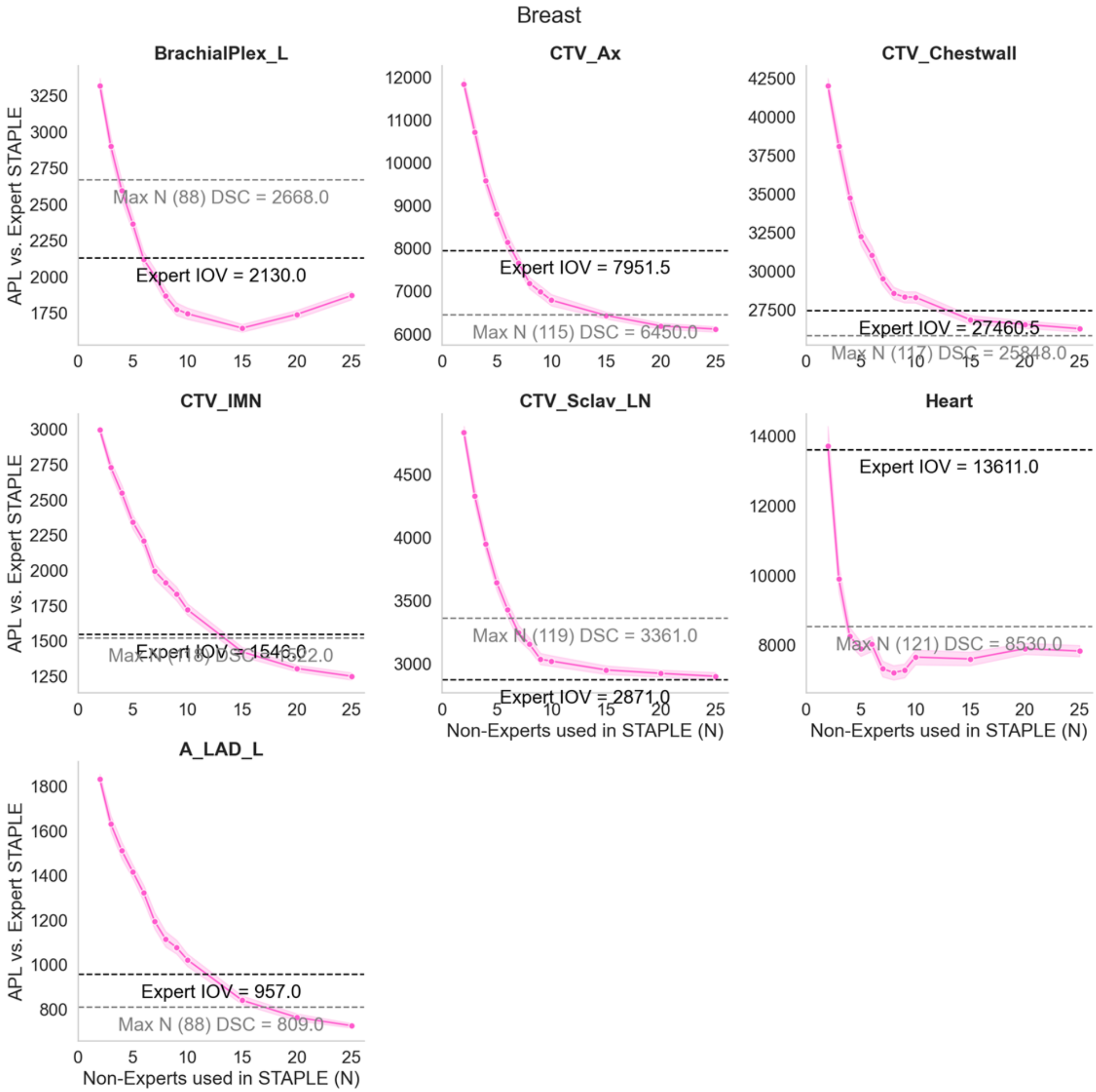


**Figure D5.** Bootstrap experiment results for H&N case using APL.

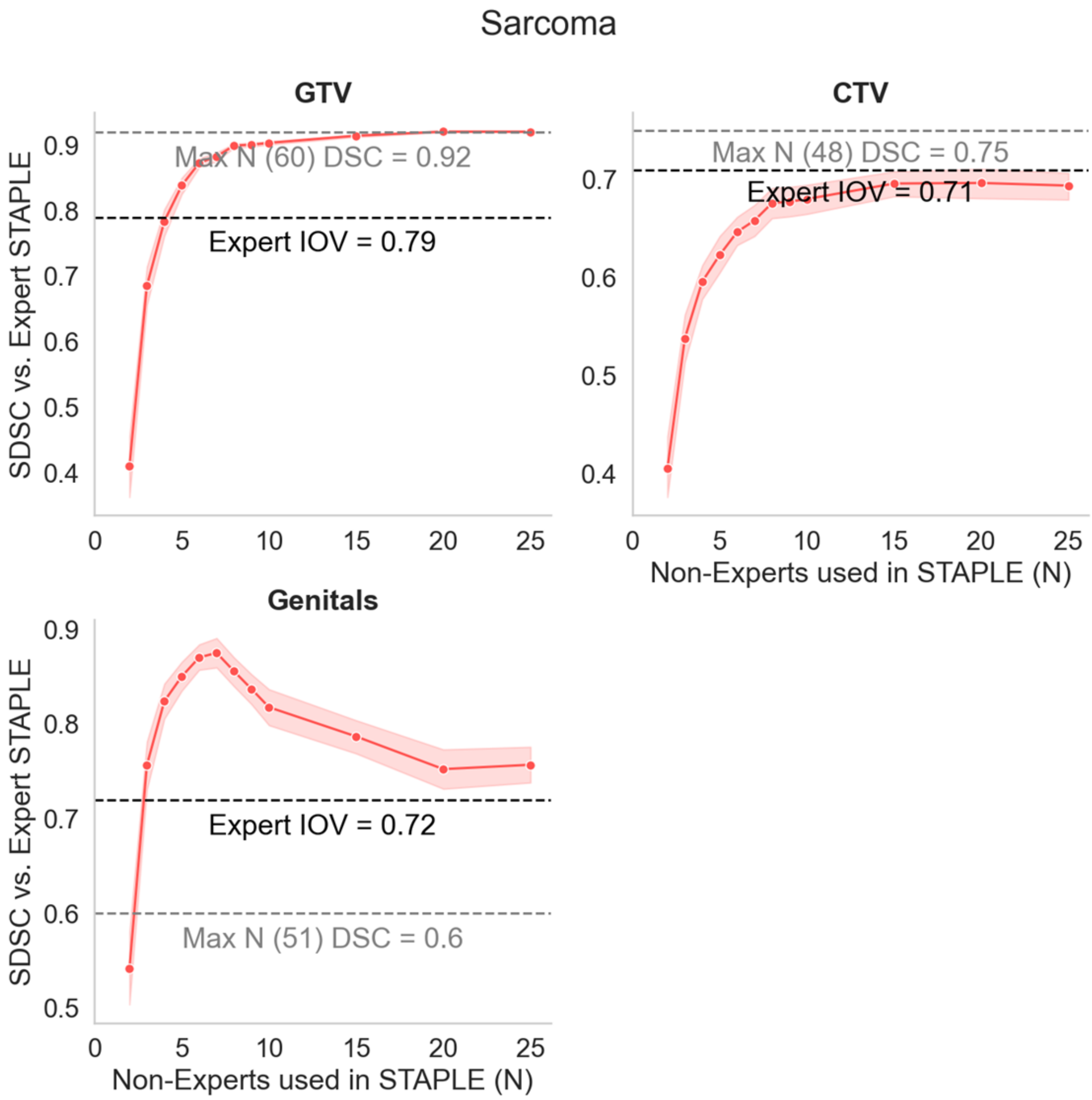


**Figure D6.** Bootstrap experiment results for Sarcoma case using SDSC.

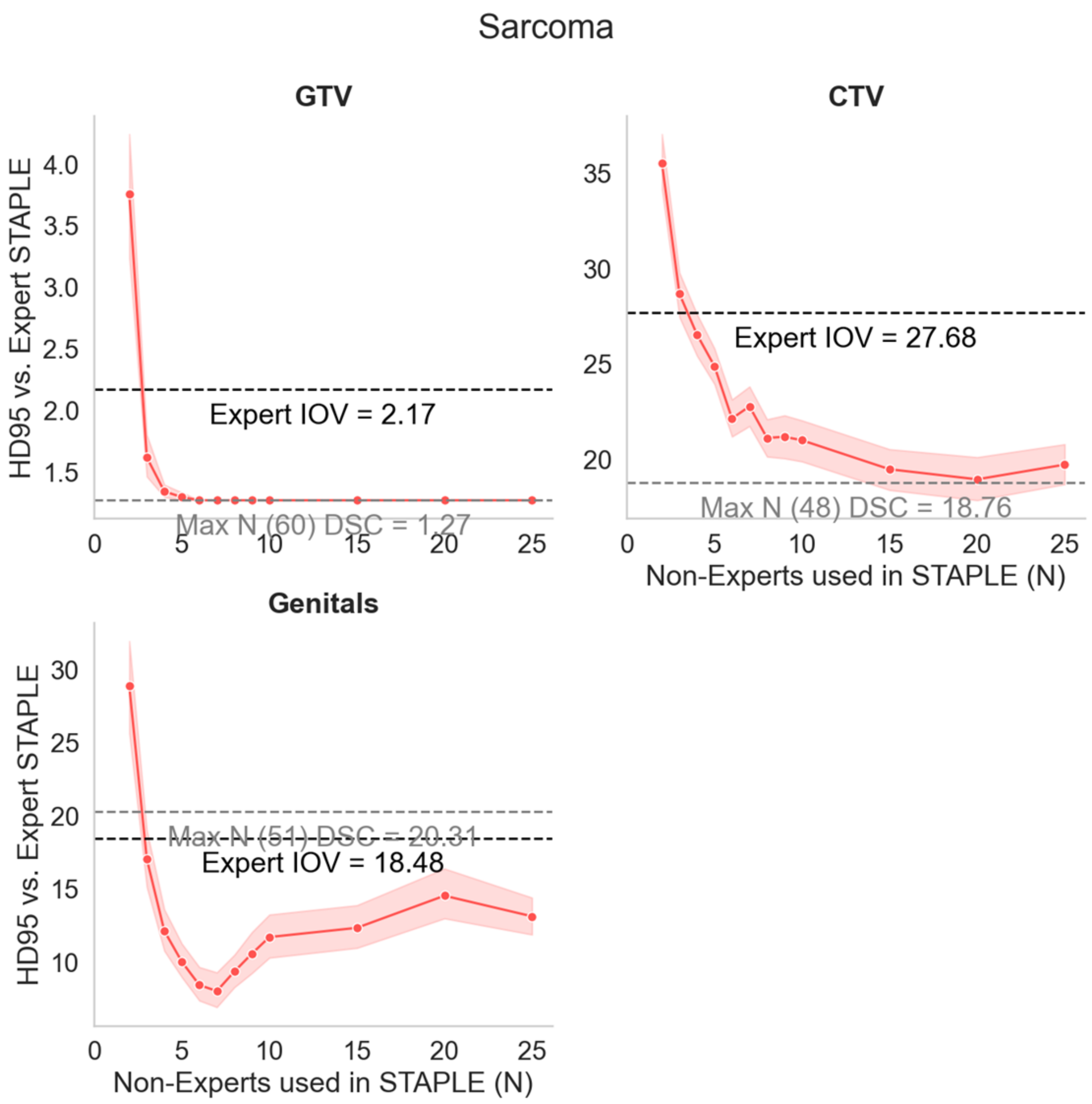


**Figure D7.** Bootstrap experiment results for Sarcoma case using HD95.

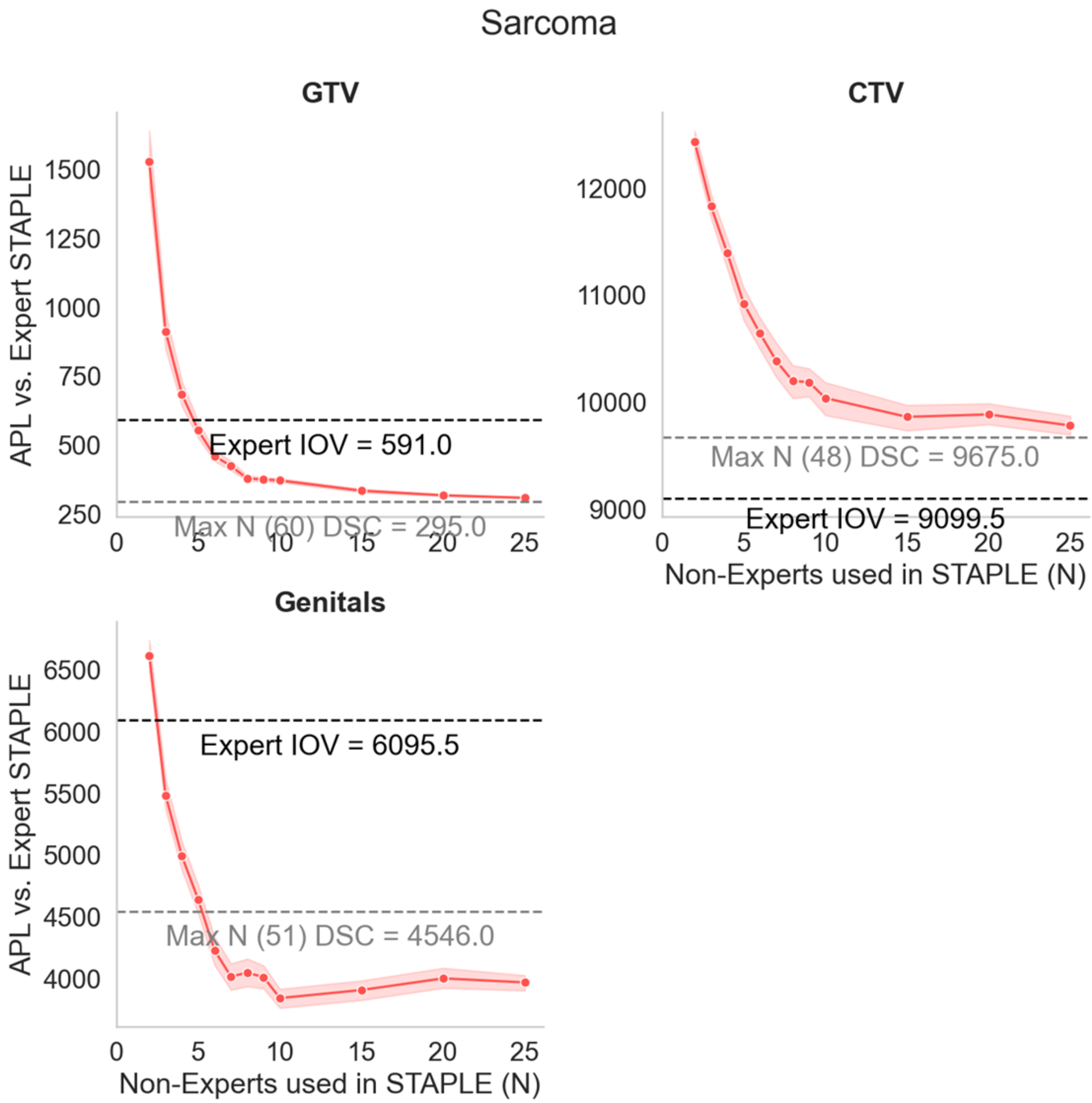


**Figure D8.** Bootstrap experiment results for Sarcoma case using APL.

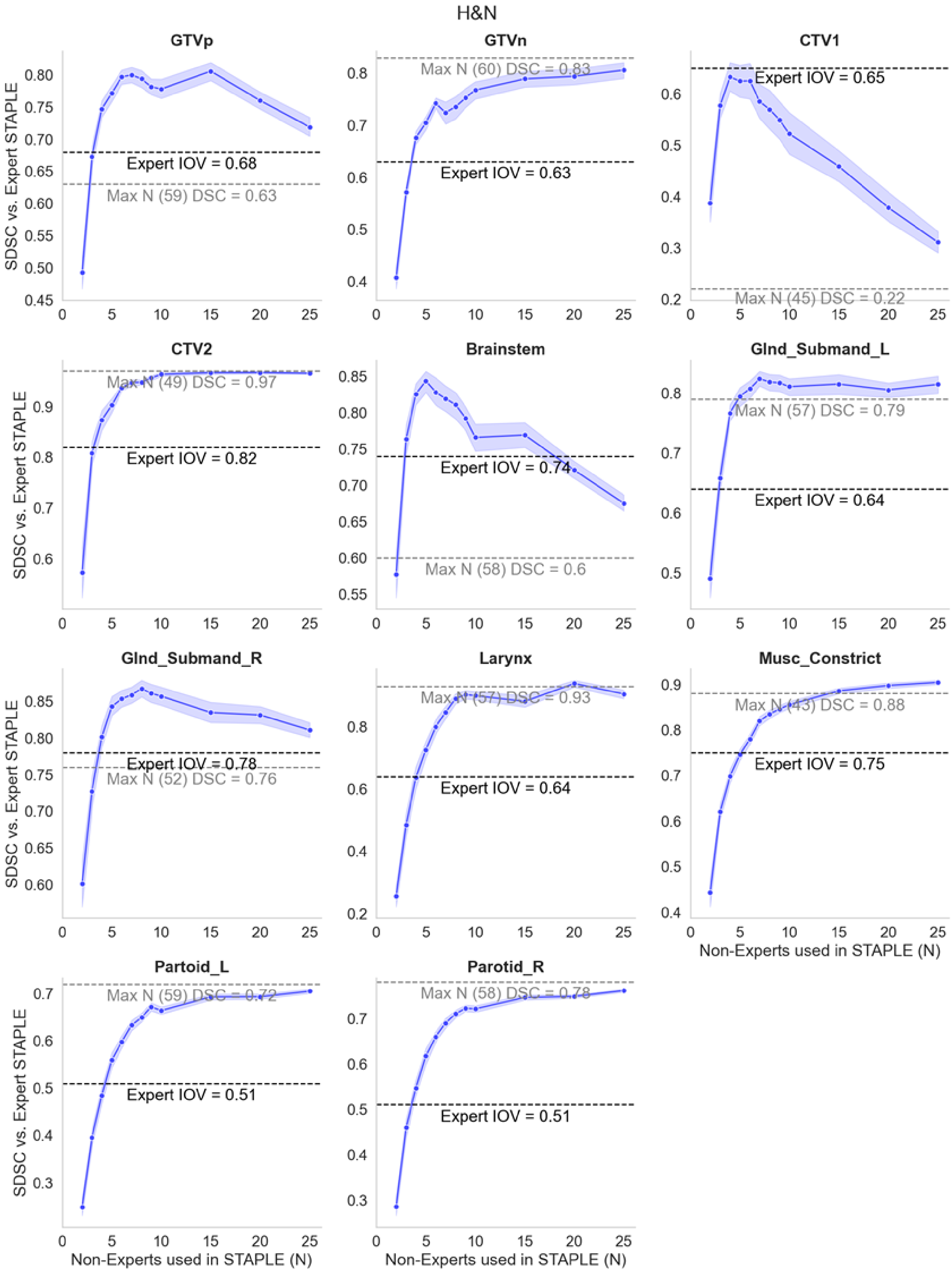


**Figure D9.** Bootstrap experiment results for H&N case using SDSC.

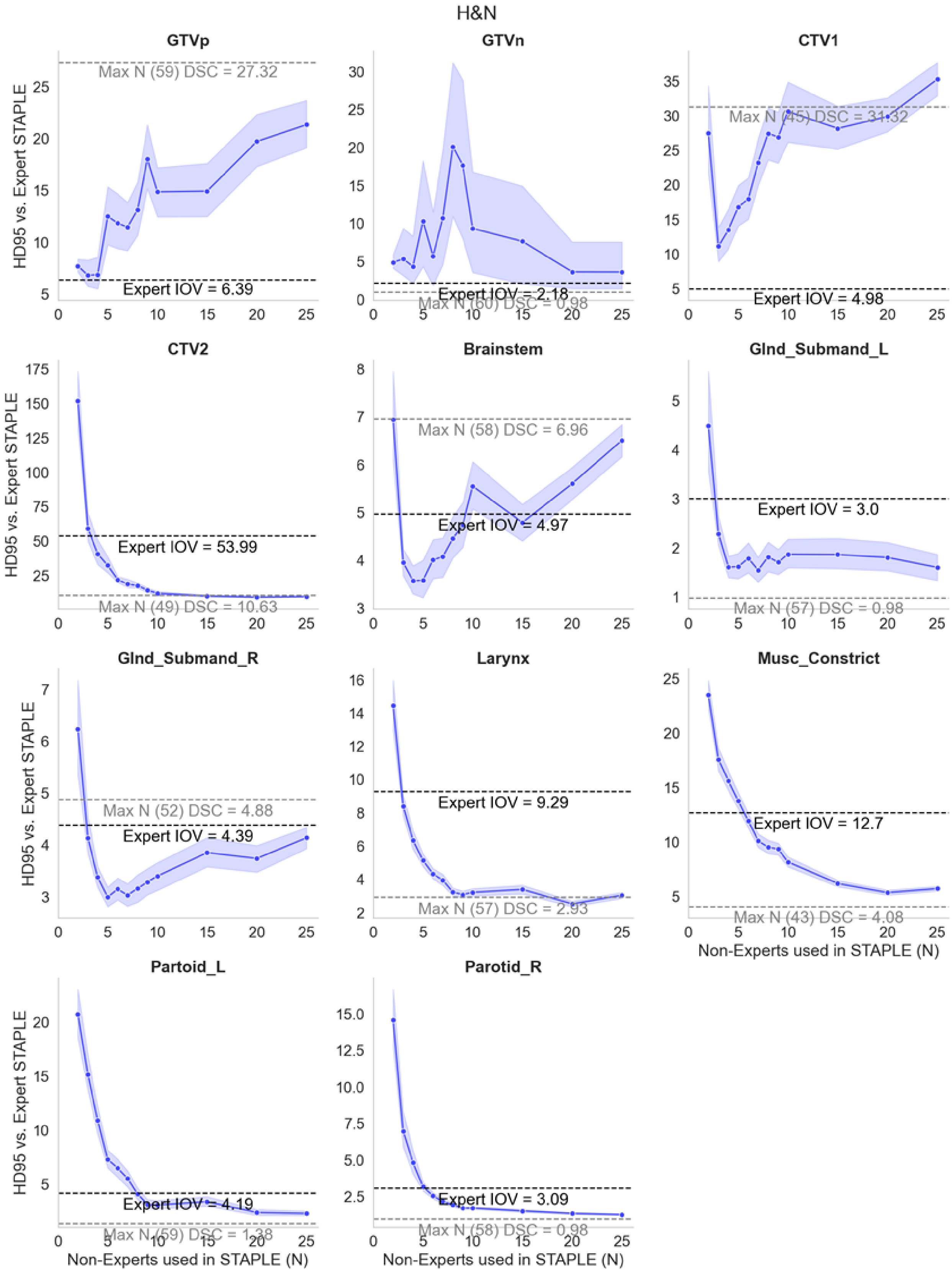


**Figure D10.** Bootstrap experiment results for H&N case using HD95.

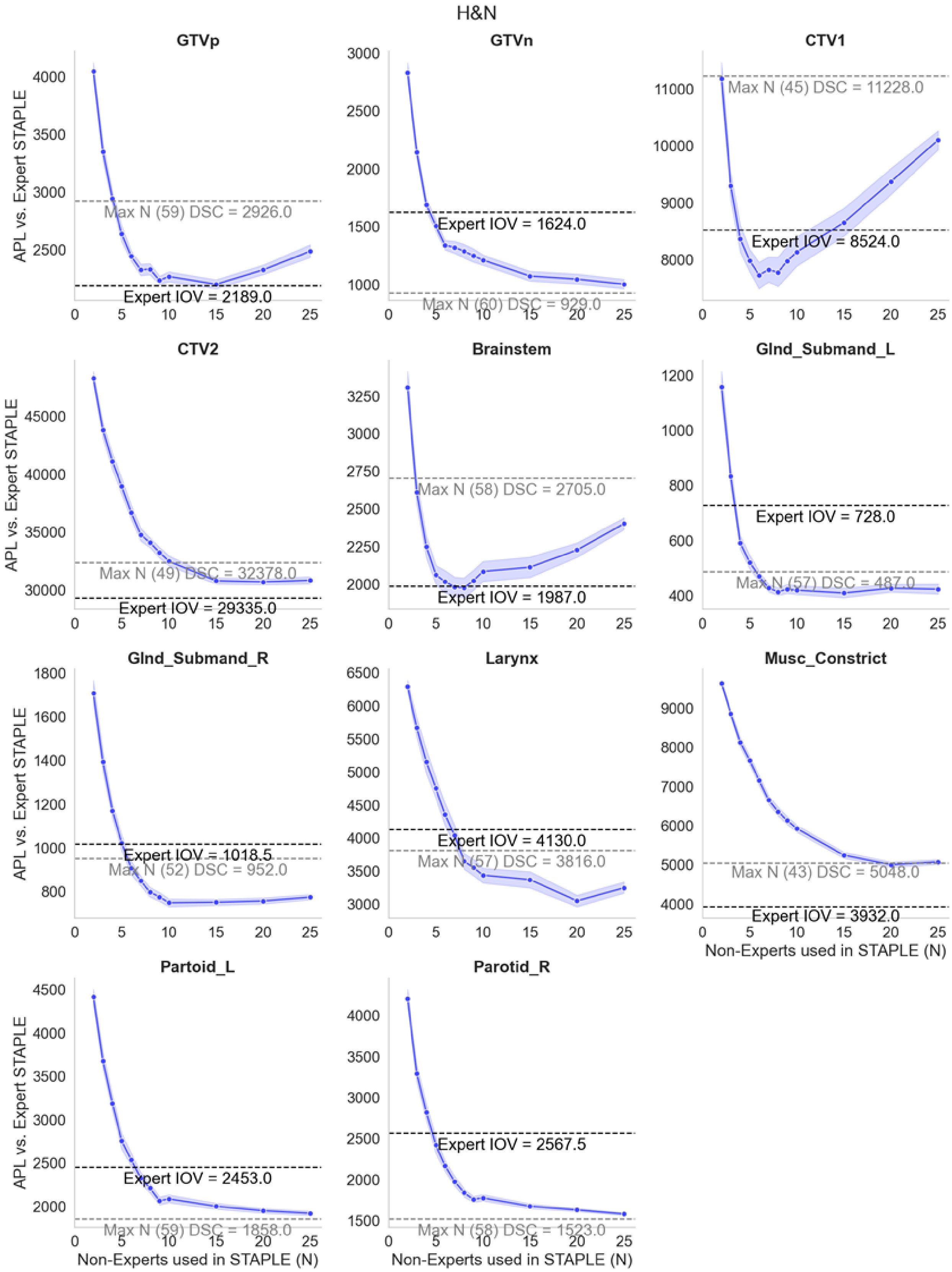


**Figure D11.** Bootstrap experiment results for H&N case using APL.

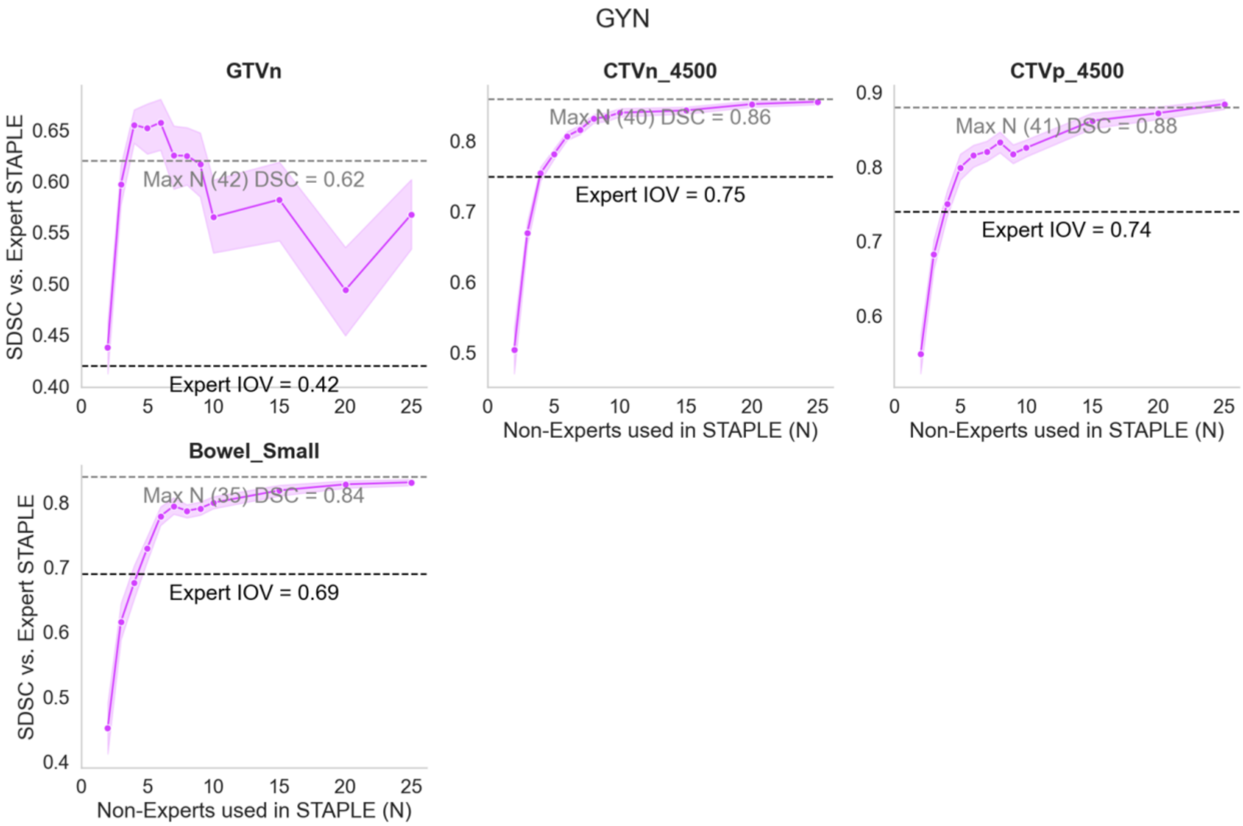


**Figure D12.** Bootstrap experiment results for GYN case using SDSC.

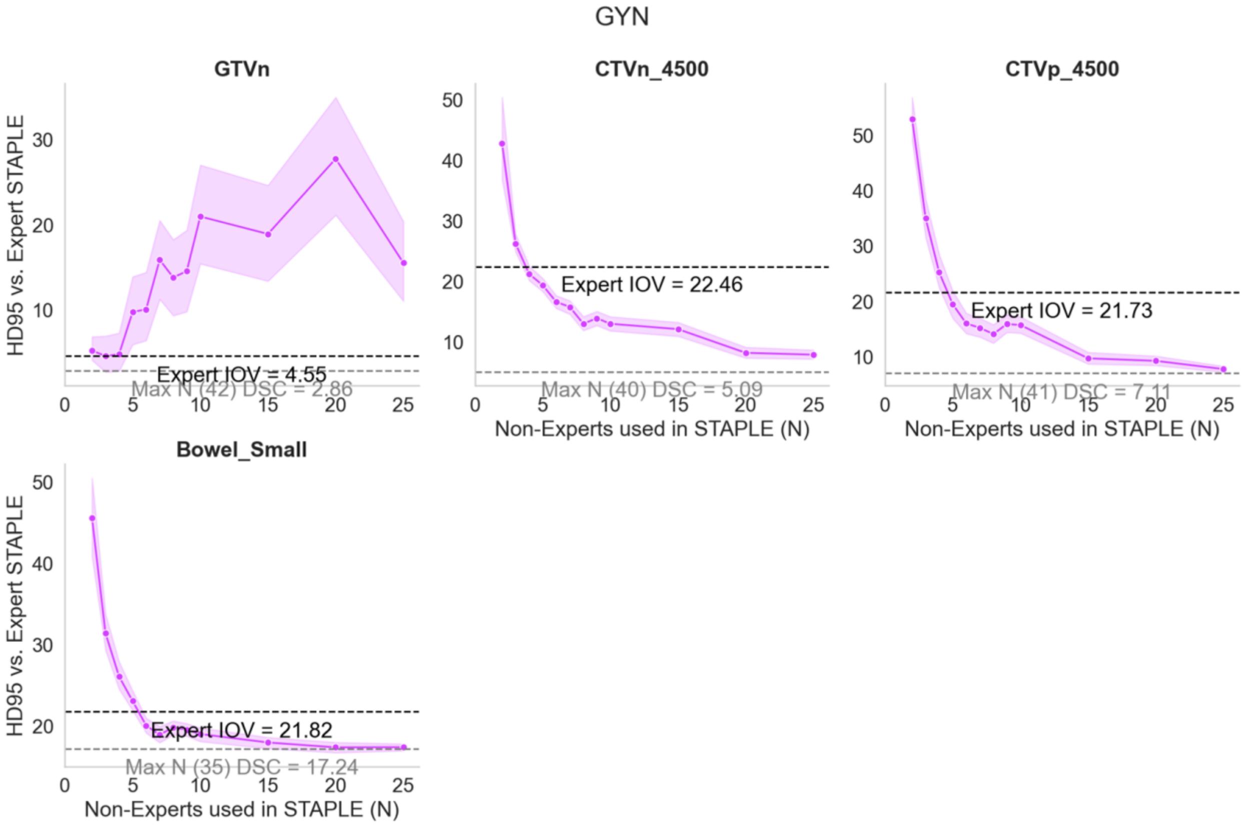


**Figure D13.** Bootstrap experiment results for GYN case using HD95.

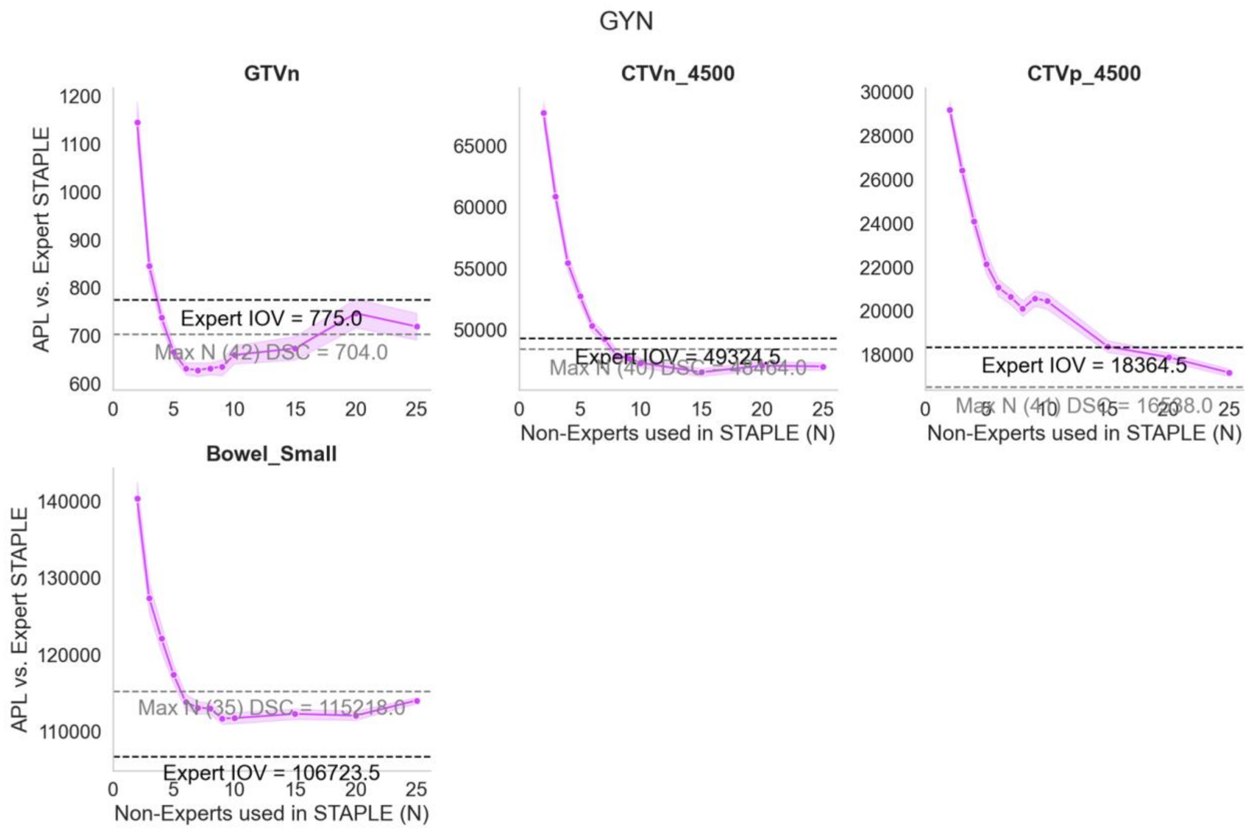


**Figure D14.** Bootstrap experiment results for GYN case using APL.

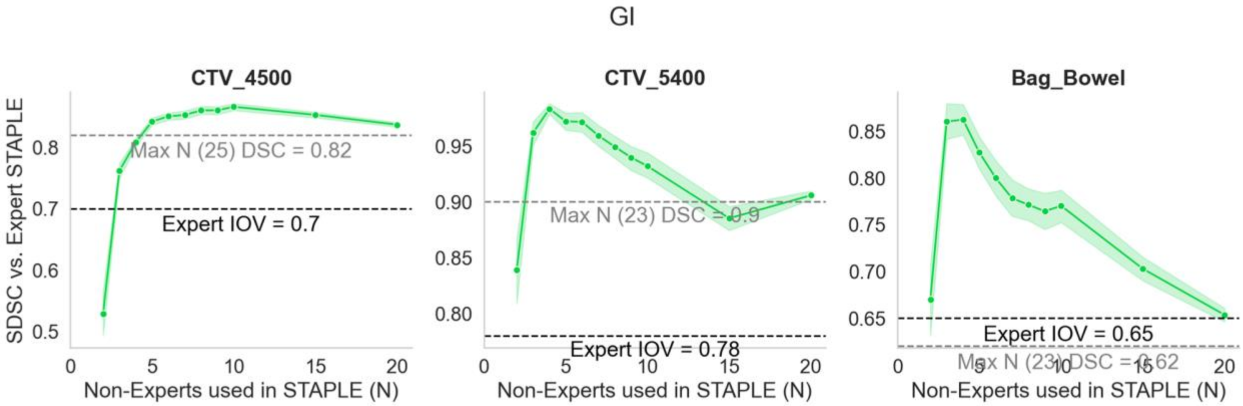


**Figure D15.** Bootstrap experiment results for GI case using SDSC.

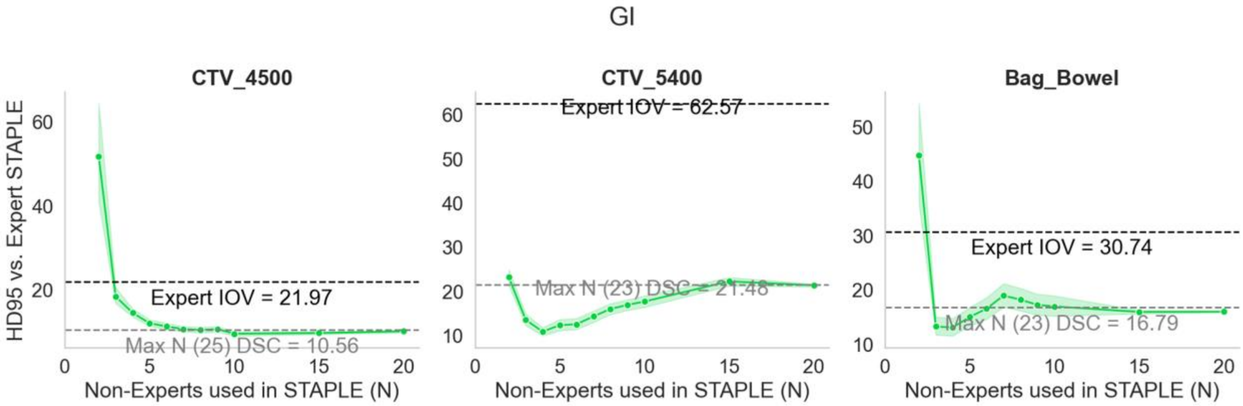


**Figure D16.** Bootstrap experiment results for GI case using HD95.

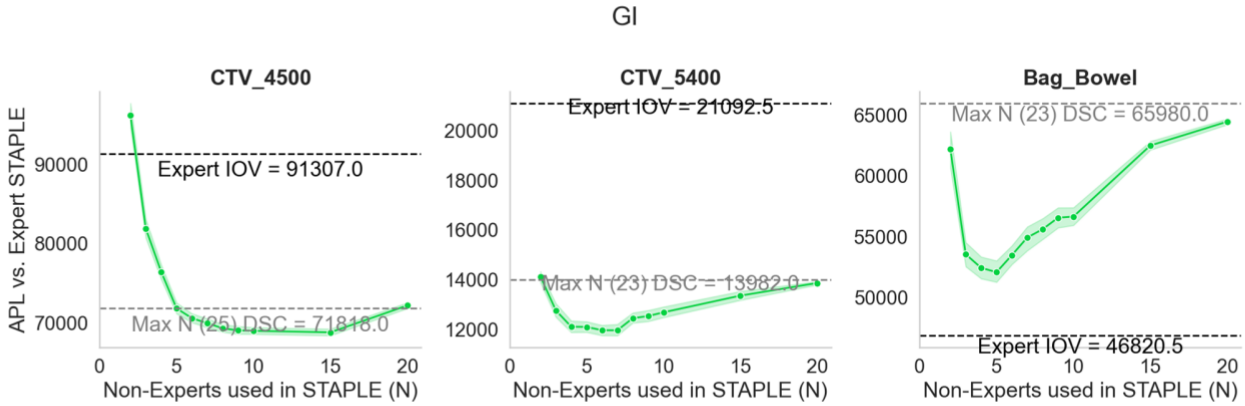


**Figure D17.** Bootstrap experiment results for GI case using APL.

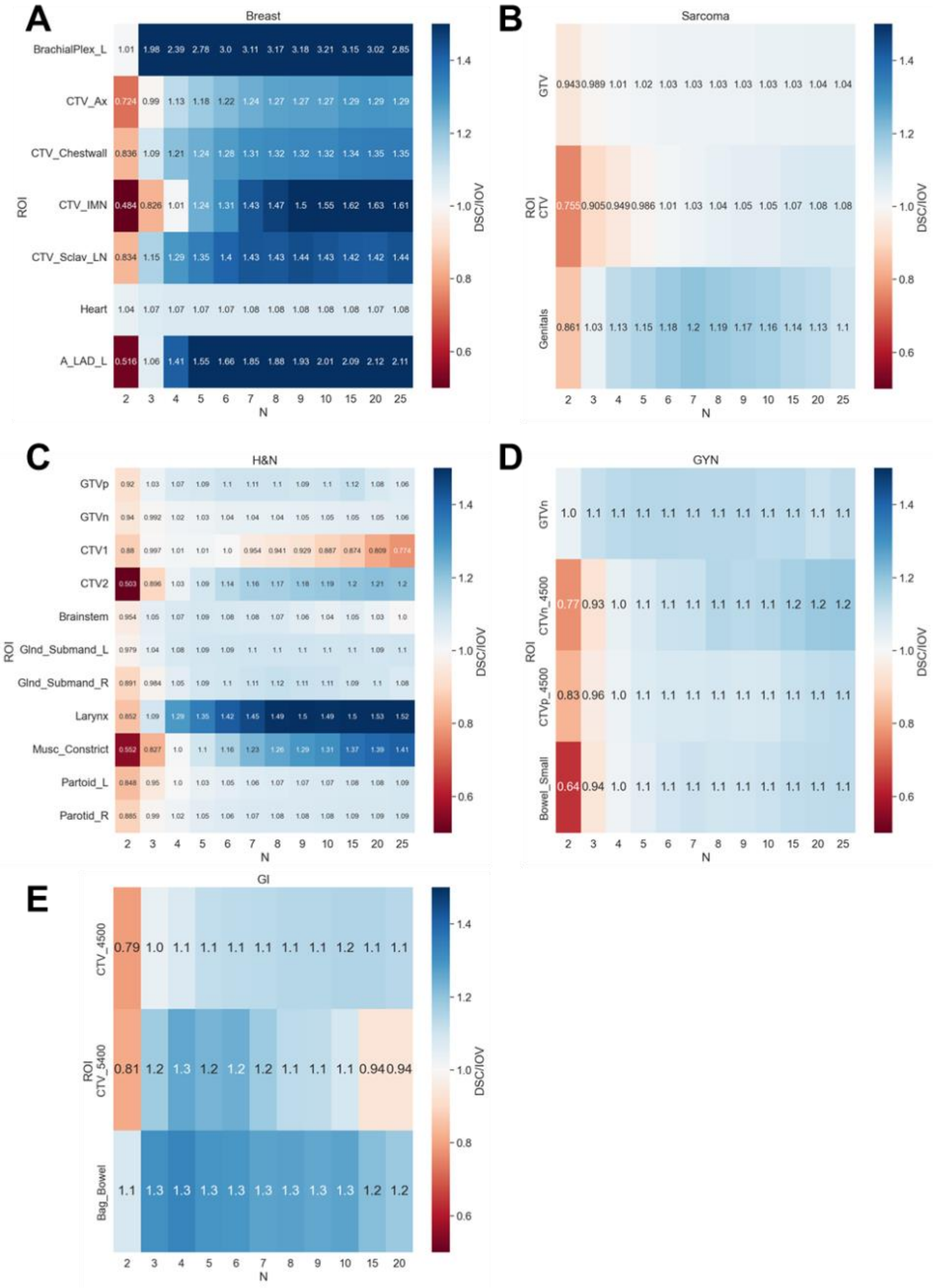


**Figure D18.** Heatmap of interobserver DSC ratio (median metric for non-expert STAPLE / median expert interobserver value) for variable number of non-expert observers used in STAPLE with respect to region of interest (ROI). The median DSC for the given non-expert subset was divided by the corresponding median expert interobserver value to yield a new measure of segmentation performance, referred to as the interobserver DSC ratio. If the interobserver DSC ratio is < 1 it is considered not clinically acceptable, if the interobserver DSC ratio is >=1 it is considered clinically acceptable. Breast, sarcoma, head and neck (H&N), gynecologic (GYN), and gastrointestinal (GI) cases are shown in (**A**), (**B**), (**C**), (**D**), and (**E**), respectively.

### Appendix E. Non-Expert vs. Expert ROI Volume Comparisons

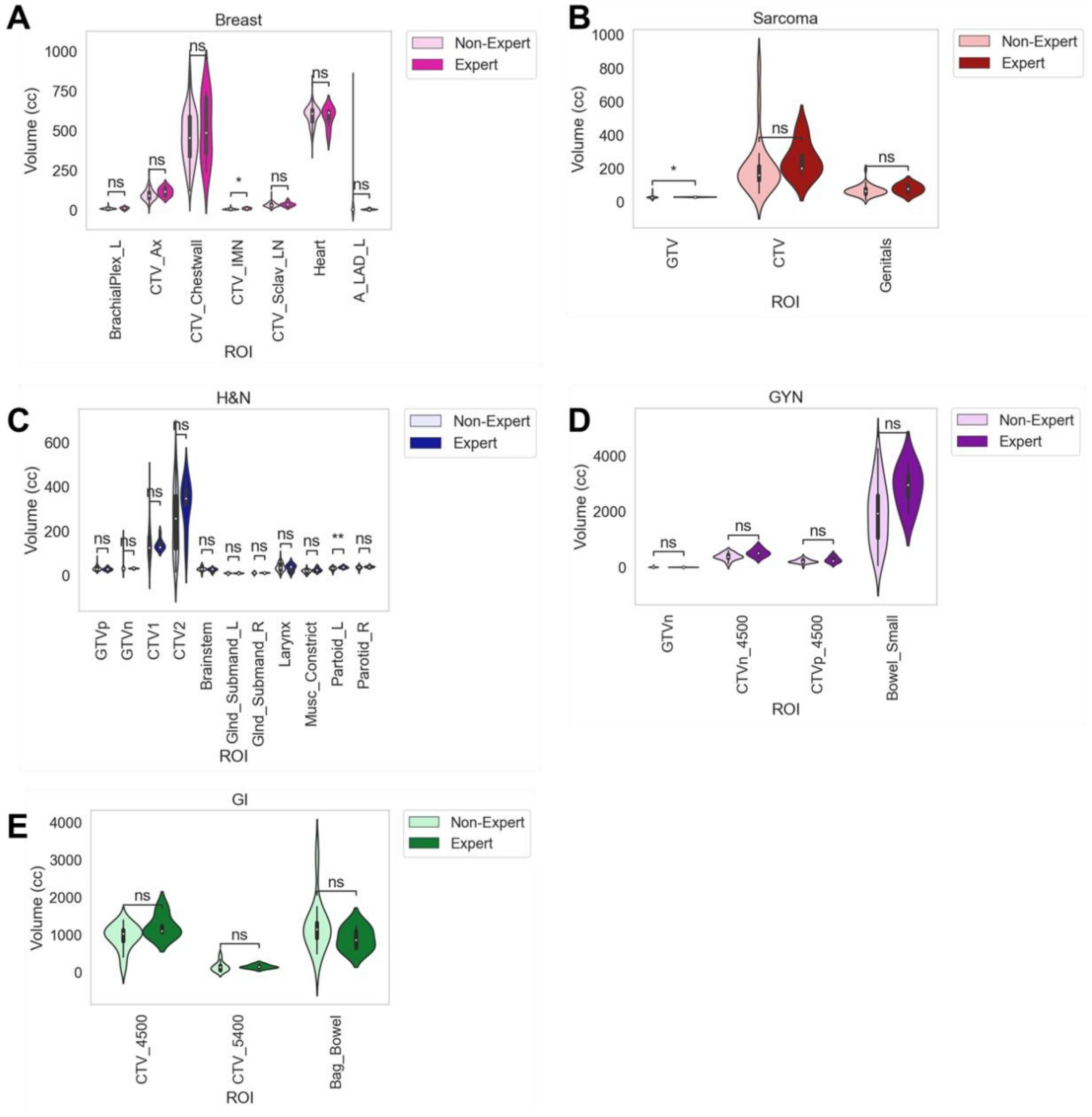


**Figure E1.** Region of interest (ROI) volume comparisons for radiation oncologists of varying expertise (non-expert, expert). Breast, sarcoma, head and neck (H&N), gynecologic (GYN), and gastrointestinal (GI) cases are shown in (**A**), (**B**), (**C**), (**D**), and (**E**), respectively. Stars above plot indicate Mann Whitney U test level of significance: ns: p <= 1.00e+00; *: 1.00e-02 < p <= 5.00e-02; **: 1.00e-03 < p <= 1.00e-02; ***: 1.00e-04 < p <= 1.00e-03; ****: p <= 1.00e-04. Abbreviations: _l = left, _r = right, gtv = gross tumor volume, ctv = clinical target volume.
